# Diagnostic analytics of routine Clinical Competency Committee data of six cohorts in family medicine program in the UAE, utilizing Milestones, EPA, and ITE

**DOI:** 10.64898/2026.07.13.26356644

**Authors:** Latifa Baynouna AlKetbi, Nico Nagelkerke, Amal Alzarouni, Mouza AlKwuiti

## Abstract

**Background:** In Competency-based medical education (CBME), longitudinal data is generated continuously. The judgments a Clinical Competency Committee (CCC) makes about trainee learning and performance are a valuable resource, supporting both resident and program development. Such data as well can enables the evaluation of rating quality and of CBME instruments such as Milestones and Entrustable Professional Activities (EPAs) which can help address a gap in the CBME literature, where evidence on the performance of these instruments remains limited.

**Objective:** Routinely gathered CCC data of six cohorts in a four-training ACGME-I-accredited family medicine residency in Al Ain, United Arab Emirates, was studied to describe growth trajectories, rating-system behavior, and the concurrent agreement of CBME instruments. As well as investigating the prospective predictive validity of two CBME instruments, EPA and Milestones, and the In-Training Exam (ITE).

**Methods:** The longitudinal CCC data for 80 residents across six cohorts (2019-20 to 2024-25) were assessed at up to eight time points (mid- and end-year; R1-R4). The pooled dataset included 10,458 EPA item ratings across 334 resident-time points, 5,021 Competency Milestone item ratings across 285 resident-time points, and 185 ITE scores.

Five research questions were examined: growth trajectories; within- and between-resident variation and straight-lining (identical scores on assessed items at a single time point); EPA-Milestones agreement; the validity of supervisor ratings against the ITE (anchoring diagnostic, same-year correlations, prospective regressions); and EPA blueprint fidelity (the mapping of EPAs against the ACGME-I subcompetency). Al Ain trajectories were benchmarked against an international family medicine reference.

**Results:** All three instruments rose steadily across the eight timepoints. By End R4, the Milestones mean (4.00, range 3.83-4.24) matched US end-of-training norms (3.84-4.02). With regards to rating quality, pooled R1-R3 Milestones straight-lining was 2.3% (EPA 0%), below US benchmarks; between-resident discrimination was preserved (SD 0.41-0.54); and longitudinal halo was ruled out (within-domain growth-slope r = 0.61 vs across-domain r = 0.37).

End R1 Overall EPA was the strongest prospective predictor of Final Competency (B = 0.96, p < 0.001) and Final ITE (B = 96.88, p = .006). Medical Knowledge ratings were independent of prior ITE scores from Mid R2 onward, and End R2 MK ratings predicted ITE 17 months later at r=0.88, confirming supervisor judgment was not anchored to test results.

With regards whether individual EPAs correlate with individual Milestone subcompetencies at each timepoint, a significant EPA and Milestones correlations were negligible at End R1 (1 of 222 item-level cells significant) and converged by End R3 (36 cells), while resident-mean stepwise regressions showed the two instruments (EPA and Milestones) behaved as overlapping predictors throughout, indicating that EPAs and Milestones are complementary at the level of specific content but convergent at the level of aggregate resident judgment.

Blueprint fidelity rose from 30% of cells reaching r ≥ 0.40 at End R2 to 80% at End R3 in the same cohort, indicating that apparent fidelity is materially affected by measurement timing.

**Conclusion:** By graduation, residents demonstrated substantial and progressive competency achievement across both instruments, with the majority reaching the entrustable threshold on both EPA and Milestone ratings. The rating system demonstrated disciplined assessment behavior of supervisors and both concurrent and prospective validity relative to the ITE. Overall EPA at End R1 was the strongest prospective predictor of all three terminal outcomes, final ITE score, graduating Competency Milestones, and graduating overall EPA, outperforming Milestones and baseline knowledge.

Routine CCC data support an evidence-based quality assurance framework spanning rater-process diagnostics, outcome-validity diagnostics, and the asymmetric-instrument diagnostic, requiring no additional data collection beyond existing program processes.

## Introduction

Competency-based medical education (CBME) requires a greater focus on comprehensive, longitudinal assessment systems that monitor learners’ progress and readiness for independent practice. A focus endorsed heavily by accreditation requirements [1] but as well motivated by concerns that time- and exposure-based training didn’t reliably ensure competence. Meeting such requirements calls for the development of efficient assessment tools to measure competence and assess the impact of this paradigm shift on physicians’ competence, [2] [3]. Evidence in the area of assessing CBME and the tools used is building, although it does not provide clear effectiveness data. A recent meta-analysis indicated improved clinical performance with competency-based curricula [4]and showed genuine learning gains. Nevertheless, research in this area remains limited amid ongoing challenges, including heterogeneity in implementation and questions about assessment methodology [5, 6].

Most ACGME and ACGME-I programs now use a Clinical Competency Committee (CCC) to integrate workplace-based ratings, examination scores, and other data into developmental judgments about each resident’s progression toward unsupervised practice [7]. CCC data, including Milestones, constitute a substantial longitudinal record of rater behavior, instrument performance, and resident progression. When analyzed systematically, these data reveal trends within and across programs and institutions and can be operationalized for ongoing program improvement [8]. Learning analytics (LA), the interpretation of multiple data sources gathered on trainees to assess progress, predict performance, and identify issues for feedback and individualized learning plans [9], extends this further by informing the CCC’s function, the Program Evaluation Committee’s review, rating-quality improvement, and timely feedback to learners. A challenge of this approach is that data management and analysis vary across programs due to differences in context, institutional culture, assessment methods, and the support tools needed to assess CCC rating quality, consistency, and fairness.

Variation among residency programs was found to be partly due to the residency program and, to a lesser degree, to learners and medical school, highlighting the influence of curricular, instructional, and programmatic factors on resident performance throughout residency [10] and calls to investigate the consistency and fairness of CCC ratings practices, mitigate uniform rating, and improve the degree to which ratings reflect individual resident competency as opposed to the residency program[11] [12] [13]. Identifying these practices may provide feedback to improve assessments and calibrate outcomes against external benchmarks.

While the ACGME mainly uses the Milestones framework, Entrustable Professional Activities (EPAs) are emerging as valuable assessment tools that provide additional rating opportunities, expand data, and stimulate reflection [14] [15]. How EPAs and Milestones interact, and whether their combined use enhances learning and CBME, remain early-stage research questions [16, 17]. Evidence shows strong correlations between EPA supervision ratings and Milestone levels across various specialties [18, 19], but the extent to which these assessments influence educational decisions remains unclear. National studies have described aggregate CBME patterns [20, 21], but program-level research is limited, and tools for evaluating rater validity, accuracy, and precision, as well as resident progression and program development over time, are scarce. Existing indicators, such as the ACGME Milestones’ straight-lining metric [20], are examples, but such quality assurance practices are still under development.

In this study, we apply learning analytics to six years of routinely collected CCC data from a family medicine residency program, which includes ITE scores, EPA ratings, and Milestone ratings. The analyses span rating-level and program-level diagnostics: growth trajectories, and rater behavior, including possible bias; concurrent agreement between EPA and Milestones; the validity of supervisor workplace ratings against the ITE; the predictive value of early ratings; and blueprint fidelity.

## Methods

### Study design and setting

This retrospective cohort study at SEHA Al Ain Clinics, UAE, is the first to apply learning analytics across three assessment tools, 19 ACGME-I Milestones, a 37-item EPA inventory, and the In-Training Examination, within a single residency over six cohorts (2019-20 to 2024-25).

### Sample and data

Eighty residents across six cohorts (C1-C6) were assessed at up to eight timepoints (mid- and end-year for R1-R4). Cohort contributions varied by instrument (Table 1): all cohorts contributed ITE data, though collection windows differed by cohort (C1: R4 only; C2: R3-R4 only; C3-C6: from R1 onward within their available training years). C1 contributed retrospective End R4 EPA data for 25 of 37 items; C2 contributed EPA data from End R3 onward (Timepoints 6-8) for the same 25 items. Both C1 and C2 are excluded from Competency Milestone analyses: C1 has no Milestone data, and C2 used a legacy 1-9 scale incompatible with the standardized 1-5 scale used by C3-C6, which formed the primary sample for Milestone and EPA analyses.

**Table 1.** Cohort overview and analytic dataset.

| Cohort | Year | n | Training status | EPA TPs | Comp TPs | ITE rounds | EPA item ratings | Comp item ratings | ITE | Notes |
| --- | --- | --- | --- | --- | --- | --- | --- | --- | --- | --- |
| C1 | 2019-20 | 10 | Graduated 2023 | TP8 only | - | R4 only | 250 | 0 | 10 | Pre-EPA system. Only retrospective End R4 EPA (25 of 37 items available). ITE at R4 only; R1-R3 not collected. |
| C2 | 2020-21 | 18 | Graduated 2024 | TP6-8 | - | R3, R4 | 1,430 | 0 | 35 | Legacy 1-9 Competency scale; excluded from 1-5 Comp analyses. EPA from End R3 onward (3 TPs). No R1/R2 ITE collected. |
| C3 | 2021-22 | 18 | Graduated 2025 | TP2-8 | TP1-8 | R1-R4 | 3,788 | 2,332 | 70 | Primary longitudinal cohort. EPA from End R1 onward (7 TPs). Full Comp coverage. 2 residents missing R3/R4 ITE. |
| C4 | 2022-23 | 15 | Currently R4 | TP1-6 | TP1-6 | R1-R3 | 2,902 | 1,592 | 41 | Through End R3 entered; TP7-8 not yet entered. 1-2 residents missing per ITE year. |
| C5 | 2023-24 | 12 | Currently R3 | TP1-4 | TP1-4 | R1, R2 | 1,584 | 846 | 22 | Through End R2 entered. 1 resident missing per ITE year. |
| C6 | 2024-25 | 7 | Currently R2 | TP1-2 | TP1-2 | R1 | 504 | 251 | 7 | Newest cohort; R1 data only. |
| Total |  | 80 | Across 6 cohorts | varies | varies | varies | 10,458 | 5,021 | 185 | C3-C6 used for 1-5 Comp analyses |

Note. TPs = assessment timepoints (TP1=Mid R1 through TP8=End R4). EPA item ratings = sum of non-missing observations across all EPA items at all timepoints per cohort. Comp item ratings = sum of non-missing observations across all 19 Milestone items at all timepoints per cohort; C1 and C2 excluded from Comp counts (C1: no Comp data; C2: legacy 1-9 scale). ITE scores = number of individual ITE administrations with non-missing scores. EPA14, 18, 22, and 28-37 were not assessed in C1 or C2, and were absent at the earliest timepoints in C3. EPA32 was absent at TP5 across all cohorts. SBP\_Q4 was introduced partway through data collection. Counts are observed (non-missing) item-level observations and reflect both missingness and differences in which EPA items were in use for a given cohort and timepoint.

The observed analytic dataset comprised 10,458 EPA item ratings (C1: 250; C2: 1,430; C3: 3,788; C4: 2,902; C5: 1,584; C6: 504), 5,021 Competency Milestone item ratings (C3: 2,332; C4: 1,592; C5: 846; C6: 251), and 185 ITE scores (C1: 10; C2: 35; C3: 70; C4: 41; C5: 22; C6: 7). Analytic sample sizes are reported with each result because cohort composition varied across timepoints.

### Instruments

#### Milestones

19 subcompetencies across six ACGME core domains: Patient Care (5 items), Medical Knowledge (2), Systems-Based Practice (4), Practice-Based Learning and Improvement (2), Professionalism (3), and Interpersonal and Communication Skills (3). The 2019 Milestones were used during data collection; the 2025 Milestones 2.0 update is not included [22]. (Appendix 1)

#### EPAs

Originally 29 items (2018-2021), mainly based on the Association of Family Medicine Residency Directors. The program’s assessment framework was based on the Association of Family Medicine Residency Directors’ Entrustable Professional Activities (EPAs) for Family Medicine [AFMRD 2015] [23], expanded to 37 EPAs by a group comprising all five UAE family medicine program directors, along with several experienced core faculty and family medicine departmental representatives, who started from the initial version and drew on international family-medicine EPA references [23, 24]. The group revised and selected items, adapted them to the UAE setting, mapped each to the six ACGME-I core competencies, and defined the accompanying entrustment rubric, all of which were approved by the group (Appendix 1). Rated 1-5. EPA14, 18, 22, and 28-37 were assessed from R2 onward in earlier cohorts. EPA32 was not assessed at Mid R3 (Timepoint 5) across any cohort.

#### ITE

Annual knowledge test scored 200-800, administered by the American Board of Family Medicine.

Both Milestones and EPAs are rated using the ACGME grid based on the Dreyfus model, with Level 4 indicating competence for unsupervised practice and Level 5 representing aspirational performance.

### Assessment process

Initial ratings are generated at each family medicine center by the assessors in the Family Medicine center who are core faculty and supervisors in the program using the program matrix (Appendix 1). Please say a few words about the composition of this group. The Clinical Competency Committee (CCC) consolidates these into consensus scores twice a year (December-January and May-June), combining EPA ratings with other data to complete the ACGME-I Milestones. Residents remain with a single Family Medicine Clinic throughout training, maintaining rater continuity across rotations and weekly clinics.

### Conceptual framework

Five research questions yielded 13 diagnostic analyses of raters and programs (Appendix 2).

Process, Rater level: Q1-Q3: Diagnostics analyze rater behavior, such as straight-lining, within- and between-resident variation, and halo effects, without relying on external criteria. These diagnostics don’t enhance rating accuracy but are valuable for rater development when shared as feedback.

Q1. Growth trajectories across training years. (a) Pooled per-instrument growth trajectories at all eight timepoints; (b) per-domain Milestones comparisons against the US benchmark; (c) between-cohort differences in EPA ratings.

Q2. Within- and between-resident variation and straight-lining. (a) Within-resident variation, (b) between-resident variation, and (c) straight-lining rate.

Q3. EPA and Milestones rating agreements. (a) Per-timepoint correlations between Overall EPA and the six Competency domains at four selected timepoints; (b) per-resident stalling (identical scores on assessed items at a single time point).

Outcome, Program level: Q4-Q5: Outcome-validity diagnostics assess program-level agreement with external benchmarks, including blueprint fidelity (EPA and subcompetency), anchoring-bias diagnosis (MK vs prior/future ITE), same-year MK and ITE correlation, and workplace predicting ITE. These are reported only at the program level to prevent raters from shifting ratings to improve validity statistics, which could bias subsequent estimates.

Q4. Validity of supervisor ratings versus the ITE. (a) Anchoring, whether MK ratings reflect independent supervisor judgment rather than test-score anchoring; (b) concurrent association between MK ratings and same-year ITE; (c) prospective prediction of terminal outcomes from early ratings.

Q5. Blueprint fidelity. (a) Sensitivity (blueprint-mapped cells with significant r) and specificity (unmapped cells correctly non-significant); (b) within-domain growth-slope correlations.

### Statistical analysis

Analyses were performed in IBM SPSS Statistics v30. All tests were two-tailed with α = 0.05. Means with 95% CIs for EPAs, Milestones subcompetencies, and ITE were computed at each of eight reporting timepoints, with per-segment change and item-level achievement (≥3.5) tabulated by timepoint. Al Ain R1-R3 trajectories were benchmarked against US Family Medicine references reconstructed from the per-subcompetency quadratic growth-curve parameters in [14], averaged within domain. Within- and between-resident SDs were computed at the item level at each timepoint, and straight-lining (zero within-resident item-level SD) was flagged at an a priori threshold of >10% at any timepoint or a monotonic upward trend. Resident-level Pearson correlations between Overall EPA and each Competency domain were summarised across timepoints using Fisher z-transformed pooled means. Retrospective anchoring of Medical Knowledge on ITE was assessed using Spearman ρ, with the diagnostic defined as the difference between ρ(future) and ρ(seen). Same-year domain-ITE and EPA-ITE correlations were computed at each training year.

Four linear regressions tested whether End-R1 workplace ratings predicted terminal Milestones competency, terminal overall EPA, and final ITE (the latest available ITE per resident). These were controlled for: ITE R2, which is considered the end-of-year 1 ITE assessment because it is conducted in October, while the academic year starts in September. The difference in R² relative to the ITE R2 baseline is reported.

Blueprint fidelity was evaluated at End-R3 (median per-cell N = 31) using the full 37 × 19 EPA × sub-competency Pearson matrix, with cells classified as Salient (r ≥ 0.40), Strong (r ≥ 0.60), or Excellent (r ≥ 0.80) per established conventions [25]; [26] and sensitivity/specificity computed per domain. Resident-level linear regression growth slopes (R1 to End-R3; n = 32 with complete coverage) were compared within-versus across-domain, with a > 0.20 gap interpreted as construct-valid differentiation.

## Results

### Q1. Growth trajectories across training years

#### Q1a. Pooled growth trajectories of EPA, Milestones, and ITE

All three instruments showed growth across the eight timepoints, but at different rates and with distinct final-year patterns. Pooled EPA and Competency means rose nearly in parallel from Mid R1 (EPA 1.61, SD 0.25; Competency 1.59, SD 0.29) through End R4. The Competency mean reached the Level 4 competence threshold at End R4 (3.95), whereas the pooled EPA mean plateaued near 3.6 in the final year (SD = 0.4). From Mid R3 onward, the pooled Competency mean exceeded the pooled EPA mean by 0.18 to 0.36 points (Figure 1).

ITE scaled scores increased across the four annual administrations, with the largest gain from R1 to R2 and progressively smaller year-over-year increases thereafter. ITE trajectories showed marked between-cohort variation, with more recent cohorts achieving higher scores at equivalent training stages (Appendix 3).

Growth velocity within-resident (change over 6-month intervals) was used to identify average slowdowns. EPA velocity slowed sharply from Mid R3 onward, with the three late-training intervals changing from +0.49 to −0.02 to +0.20. Milestones velocity also slowed but remained consistently positive across the same intervals (+0.36, +0.13, +0.30). Figures 2-A and 3 and Figure 4

#### Q1b. Per-domain trajectories vs. the US Milestones benchmark

At the US graduation point (End R3, US End-PGY-3, TP6), the Al Ain End R3 pooled Competency mean was 3.46, below the US mean of 3.94. By the end of R4, Al Ain’s mean (4) matched the US end-of-training range reported by Park et al. (3.84-4.02). At End R4, three domains were within ±0.10 of the US mean: PC (3.89 vs 3.98), SBP (3.83 vs 3.93), and ICS (3.92 vs 4.02). Three domains exceeded the US mean: PROF (4.24 vs 3.90), PBLI (4.09 vs 3.84), and MK (4.06 vs 3.92). US data do not extend beyond End-PGY-3, so the R4 catch-up could not be directly benchmarked against US residents at the same stage. Competency data at End R4 are based on the 2021-22 C3 cohort, the only group with final standardized scores. Appendix 4

The mean per-item between-resident SD was stable across all eight rating periods for both instruments, averaging 0.45 for EPA and 0.49 for Competency, with ranges of (0.41-0.47) and (0.43-0.54), respectively. The trajectories tracked closely, with dips at Mid-R3 (EPA 0.44; Competency 0.43) and a small rise afterward (Competency reaching 0.54 at Mid-R4 and End-R4). The lack of collapse (of EPA and competency) to zero and similar ranges suggests that rater discrimination was maintained, indicating avoidance of straight lining or central-tendency bias. Figure 2D

Per-item growth between End-R3 and End-R4 varied widely across the 37 EPAs. The slowest-growing EPAs were Adaptability/resilience (EPA32), End-of-life and palliative care (EPA12), Professional identity (EPA25), and Recovery and function (EPA5). The fastest-growing clusters in R4-specific curriculum areas were Teaching and supervising learners (EPA31), Urgent/emergent care recognition (EPA14), Management plans (EPA33), and Difficult conversations (EPA36). At the cluster level, R4-curriculum-heavy clusters, including procedural skills, managing acute conditions, and administrative/special contexts, showed mean changes of ≥0.43 units, whereas the scholarship cluster showed a smaller change of 0.21 units. Appendix 5

#### Q1c. Between-cohort differences in EPA ratings

Mean EPA ratings differed substantially between cohorts at shared timepoints, whereas Competency ratings showed smaller between-cohort differences. Among three cohorts with End R4 EPA data, the mean Overall EPA was 3.18 (C1 2019-20), 3.51 (C2 2020-21), and 3.90 (C3 2021-22), a 0.72-point range. At End R3, where C2 and C3 had EPA data, the means were 2.99 (C2) and 3.49 (C3), a 0.50-point gap. Competency ratings at End R3 for C3 and C4 were nearly identical (3.48 vs 3.44), a 0.04 gap; at End R2, with C3, C4, and C5 data, the mean spread was 0.23 points. The same residents in the same program, therefore, showed a marked difference on the EPA between-cohort but virtually none on the Competency instrument. Figure 2e

During C1-C3, the program’s EPA inventory expanded from 29 to 37 items in 2022. Per-cohort Overall EPA trajectories are shown in Figure 1; per-cohort, per-domain Milestones trajectories are shown in Appendix 3.

### Q2. Within- and between-resident variation and rate of straight-lining

The variation within and between residents’ assessments is shown in Figures 2A and 2B, and the rate of straight-lining is shown in Figure 2C.

**Q2a. Within-resident standard deviation** (across items within a single resident at a single time point), from R1 through EPA training, remained stable, with no overall change (0.37-0.48 across TP1-TP8). For Competency, variation declined, with less variation observed. It went from a 0.45-point variation in ratings given to a resident at Mid R1 to more consistent ratings, with only a 0.22-point variation by Mid R4. Figure 2A

**Q2b. Between-resident standard deviation** (spread across residents for a given EPA or Competency) remained stable on both instruments (0.41-0.54), indicating that ratings for a given resident were consistent and that ratings continued to distinguish among residents, even as within-resident scores varied over the training period. Figure 2B

#### Q2c. Straight-lining

EPA straight-lining was 0% at every time point. Milestones straight-lining remained low throughout R1-R3 (range 0-6.3%; pooled rate 2.3%). Rates rose in the graduation year to 25.0% at Mid R4 and 18.8% at End R4 (n = 16 at both timepoints), though the 95% confidence intervals were wide given the small cohort (10.2-49.5% and 6.6-43.0%, respectively). Figure 3

#### Q2d.Longitudinal halo

Overall, evidence of a longitudinal halo, the possibility that raters were affected by a global resident impression, was tested using within-timepoint correlations across domains and trajectory-level across-domain growth correlations. The across-domain growth-slope correlation was r = 0.37, while the within-domain growth-slope correlation was r = 0.61 (strong). This pattern is sufficient to decrease the likelihood of the halo effect, even though residents’ growth in other domains is correlated with their scores at any given time point.

### Q3. EPA-Milestones ratings/performance match

The resident-level Pearson correlation between mean EPA score and mean Competency score (computed at each of the eight timepoints) rose steadily from Mid R1 onwards and peaked at End R3 (r = 0.74), then declined modestly at End R4 (r = 0.74 at Mid R4, r = 0.53 at End R4). The R4 decline could at least partly be a sampling effect: with n = 16 at End R4.

An item-level heatmap of point-biserial correlations between individual EPAs and the six Competency domains at four timepoints (Appendix 6) showed the same pattern: 1 significant cell at End R1 (TP2), 18 at End R2 (TP4), 36 at End R3 (TP6, the peak), and 19 at End R4 (TP8). Per-resident stalling (proportion of items with no advance between R3 and R4) was common on both instruments; approximately 27% of EPAs and 30% of Competency items did not advance per resident, but were only weakly correlated between instruments (r = 0.29, not significant). Stalling was therefore item-specific or task-specific, not a resident-level plateau phenomenon of near-ceiling mechanisms.

### Q4. Validity of supervisor EPA/Competencies ratings against ITE

#### Q4a. Assessor-anchoring diagnostic

Because the ITE is administered annually each autumn and results are released before the mid-year (January) competency milestones rating round, MK competency could, in principle, be anchored to a recent test score rather than reflecting independent observation. To explore this, Spearman’s ρ was computed between MK ratings at each of the eight rating time points and each of the four annual ITE administrations (columns), yielding 32 correlations. Each correlation was classified by its temporal status: an ITE was “already seen” if released to supervisors before the MK rating, or “not yet administered” if the test had not yet occurred (Figure 5). Under anchoring, ρ with already-seen ITEs should dominate; under genuine independent knowledge judgment, ρ with future ITEs should be at least as high as with seen ITEs. Three temporal phases emerged. At Mid R1 and End R1 (Year 1), supervisors had observed residents for only four months and had recently seen the ITE scores (ITE R1). Mean ρ with the seen ITE (R1) at these two timepoints was 0.69 and 0.66, exceeding the mean ρ with the three future ITEs (0.45 and 0.41, respectively; difference around −0.24). This pattern is consistent with mild rater anchoring on the ITE scores during the first six months, when direct observation is limited, and the recently released test result is salient. From Mid R2 onward, the pattern reversed. As shown in Figure 5, future-ITE correlations exceeded seen-ITE correlations (difference ranged from +0.08 at Mid R3 to +0.21 at End R3). At End R2, MK ratings based only on ITE R1 and ITE R2 correlated with upcoming ITE R3 and ITE R4, with a mean ρ = 0.73, compared to a seen-ITE correlation of 0.54 (+0.19). The strongest was End R2 MK with future ITE R4 (ρ = 0.88), 17 months before the test. Since supervisors couldn’t have seen ITE R4 at the time, this forward prediction isn’t due to anchoring on past scores. This rules out anchoring as an alternative explanation for the MK-ITE concurrent and prospective correlations reported in Q4b and Q4c.

#### Q4b. Same-year correlations

Resident-level Pearson correlations between EPA/Competencies ratings and same-year ITE were computed (Table 2C). Medical Knowledge was the only domain to reach significance every year (r = 0.47-0.74, p < .05). Overall, EPA was significant from End R3 onwards (r = 0.55-0.62, p < .001). The other domains (Patient Care, Systems-Based Practice, Practice-Based Learning, Interpersonal/Communication Skills) were intermittently significant, with three to five significant points out of eight. Professionalism was the weakest, significant in only two of eight cells.

**Table 2.** Per-domain milestone benchmarking, anchoring diagnostic, concurrent validity, and prospective prediction (including instrument-independence regressions)

**2A. Concurrent workplace-rating and ITE Pearson correlations, by training year**
| Training-year rating | PC | MK | SBP | PBLI | PROF | ICS | Overall EPA |
| --- | --- | --- | --- | --- | --- | --- | --- |
| R1 - Mid (ite.r1) | +.41**<br>(49) | +.65***<br>(49) | +.33*<br>(49) | +.20 (49) | +.01 (48) | +.16 (49) | - .34 (32) |
| R1 - End (ite.r1) | +.37**<br>(49) | +.61***<br>(49) | +.30*<br>(49) | +.24 (49) | +.35*<br>(48) | +.19 (49) | +.22 (50) |
| R2 - Mid (ite.r2) | +.30 (41) | +.53***<br>(41) | +.31 (41) | +.38*<br>(41) | +.08 (40) | +.16 (41) | +.24 (43) |
| R2 - End (ite.r2) | +.18 (42) | +.47**<br>(42) | +.06 (42) | +.27 (42) | +.08 (41) | - .04 (42) | +.02 (43) |
| R3 - Mid (ite.r3) | +.50**<br>(29) | +.71***<br>(29) | +.19 (29) | +.50**<br>(29) | +.29 (28) | +.39*<br>(29) | +.23 (30) |
| R3 - End (ite.r3) | +.54**<br>(29) | +.72***<br>(29) | +.54**<br>(29) | +.51**<br>(29) | +.40*<br>(28) | +.41*<br>(29) | +.55***<br>(47) |
| R4 - Mid (ite.r4) | +.38 (16) | +.54* (16) | +.46 (16) | +.58*<br>(16) | +.41 (15) | +.41 (15) | +.62***<br>(35) |
| R4 - End (ite.r4) | +.62**<br>(16) | +.74**<br>(16) | +.60*<br>(16) | +.25 (16) | +.35 (16) | +.61*<br>(16) | +.59***<br>(45) |

**2B. Concurrent Overall EPA and Competency-domain Pearson correlations, per timepoint (N = 80; cell: r (n))**
| Timepoint | PC | MK | SBP | PBLI | PROF | ICS | Pooled (mean) |
| --- | --- | --- | --- | --- | --- | --- | --- |
| Mid R1 | - .03 (33) | - .41* (33) | - .20 (33) | - .02 (33) | - .18 (32) | - .22 (33) | - .18 (32) |
| End R1 | +.30* (50) | +.49***<br>(50) | +.42**<br>(50) | +.46***<br>(50) | +.53***<br>(49) | +.40**<br>(50) | +.44***<br>(50) |
| Mid R2 | +.72***<br>(41) | +.62***<br>(41) | +.78***<br>(41) | +.60***<br>(41) | +.64***<br>(40) | +.65***<br>(41) | +.67***<br>(40) |
| End R2 | +.55***<br>(43) | +.51***<br>(43) | +.46**<br>(43) | +.36* (43) | +.59***<br>(42) | +.67***<br>(43) | +.53***<br>(42) |
| Mid R3 | +.65***<br>(32) | +.62***<br>(32) | +.56***<br>(32) | +.54** (32) | +.70***<br>(31) | +.73***<br>(32) | +.64***<br>(31) |
| End R3 | +.71***<br>(31) | +.72***<br>(31) | +.75***<br>(31) | +.66***<br>(31) | +.77***<br>(30) | +.83***<br>(31) | +.74***<br>(30) |
| Mid R4 | +.72**<br>(16) | +.70**<br>(16) | +.74**<br>(16) | +.66** (16) | +.79***<br>(15) | +.79***<br>(15) | +.74***<br>(15) |
| End R4 | +.46 (16) | +.49 (16) | +.51* (16) | +.23 (16) | +.45 (16) | +.85***<br>(16) | +.53***<br>(16) |

2C. Prospective prediction of terminal outcomes - Models 5a (Final Comp) and 5b (Final Overall EPA)
| Predictors | Model 5a - DV: Final Overall Comp (n = 30) |  |  | Model 5b - DV: Final Overall EPA (n = 31) |  |  |
| --- | --- | --- | --- | --- | --- | --- |
|  | B | 95% CI | p | B | 95% CI | p |
| Overall Comp at End R1 (ALL_COMP_MEAN.2) | - |  |  | 0.510 | [0.087, 0.934] | .020 |
| Overall EPA at End R1 (ALL_EPA_MEAN.2) | 0.956 | [0.49, 1.42] | <.001 | 0.606 | [0.144, 1.068] | 0.012 |
| ITE R2 (control) | 0.0000 | [- 0.0017, 0.0018] | .964 | 0.0004 | [- 0.0015, 0.0022] | .689 |
| Constant | B = 1.653<br>(SE 0.488) |  |  | B = 2.441<br>(SE 0.483) |  |  |

2D. Single-predictor stepwise regressions for instrument independence - Models 5c–5e (DV: Final Overall Comp; n = 30)
| Model | Predictor (timepoint) | B | 95% CI | p | Cross-instrument predictor at same TP ($\beta$, p) | ITE R2 ($\beta$, p) |
| --- | --- | --- | --- | --- | --- | --- |
| 5c | End R1 EPA (End R1 mean) | 0.959 | [0.523, 1.396] | <.001 | End R1 Comp: $\beta$ = .178, p = .322 | $\beta$ = .007, p = .964 |
| 5d | End R1 Comp (End R1 mean) | 0.633 | [0.201, 1.065] | .006 | End R1 EPA: not entered (Model B) | $\beta$ = .138, p = .415 |
| 5e | End R2 Comp (End R2 mean) | 0.846 | [0.414, 1.279] | <.001 | End R2 EPA: $\beta$ = .208, p = .331 | $\beta$ = - .099, p = .565 |

**Table 3.** Within-domain and across-domain coherence of subcompetency growth slopes.

| Domain | Within-domain r<br>(mean across<br>pairs) | Across-domain r (vs each other domain) |  |  |  |  |  |
| --- | --- | --- | --- | --- | --- | --- | --- |
|  |  | PC | MK | SBP | PBLI | PROF | ICS |
| **Professionalism (PROF)**<br>k=3, 3 pairs | **0.79** | 0.38 | 0.37 | 0.34 | 0.32 | 0.36 avg | 0.40 |
| **Interpersonal/Comm. (ICS)**<br>k=3, 3 pairs | **0.71** | 0.42 | 0.41 | 0.38 | 0.35 | 0.40 | 0.43 |
| **Medical Knowledge (MK)**<br>k=2, 1 pair | **0.69** | 0.39 | 0.38<br>avg | 0.35 | 0.33 | 0.37 | 0.41 |
| **Practice-Based Learning (PBLI)**<br>k=2, 1 pair | **0.61** | 0.34 | 0.33 | 0.31 | 0.28 avg | 0.32 | 0.35 |
| **Systems-Based Practice (SBP)**<br>k=4, 6 pairs | **0.61** | 0.36 | 0.35 | 0.33 avg | 0.31 | 0.34 | 0.38 |
| **Patient Care (PC)**<br>k=5, 10 pairs | **0.51** | 0.40 | 0.39 | 0.36 | 0.34 | 0.38 | 0.42 |

Across-domain row-averages: PC 0.40, MK 0.38, SBP 0.33, PBLI 0.28, PROF 0.36, ICS 0.43. Every within-domain r exceeds its corresponding across-domain row-average. PC has the smallest within-across gap (0.51 vs 0.40, +0.11) and the lowest within-domain coherence; PROF has the largest gap (0.79 vs 0.36, +0.43) and the highest within-domain coherence.

Resident-level correlations between overall EPA and each of the six Competency domains rose from weak or absent at Mid R1 to consistently strong from End R2 onwards, peaking at End R3 (all six domains r = 0.66-0.83; all p < .001).

#### Q4c. Prospective prediction, early ratings as predictors of terminal outcomes

##### EPA or Competencies prediction (Models 5a, 5b)

Two stepwise linear regressions were used to test whether graduation (R4) or near-graduation (R3) Competency or EPA scores, two to three years later, were predicted by baseline ITE or early EPA/Competencies ratings (Table 2). Overall, EPA at End R1 strongly predicted graduating overall Competencies (B=0.96 [0.49, 1.42], p<.001): one unit increase in early EPA (observed range 1.43-2.94) predicted nearly one unit increase in graduating Competency (observed range 2.24-4.53). Secondly, overall Competencies at End R1 predicted graduating overall EPA (B=0.510 [0.087, 0.934], p=.020). Baseline ITE (R1 or R2) was not significant in either model (both p≥0.69), indicating that baseline knowledge-test performance did not independently improve the prediction of terminal EPA/Competencies outcomes once the cross-instrument EPA/Competencies predictor was included in the model.

##### Final (R3 or R4) ITE prediction (Models 5f, 5g)

Using stepwise linear regressions with the latest R3 or R4 as the dependent variable and all early ITE EPAs and Competencies as independent variables (Table 4), only ITE R2 was a significant predictor in both models. In addition, in Model 5f MK at End R1 contributed beyond ITE R2 (B = 55.01, p = .037), 55 scaled ITE points per one-unit increase in MK rating. Model 5g showed that Overall EPA at End R1 contributed more strongly (B = 96.88, p = .006), 97 scaled ITE points per one-unit increase in Overall EPA rating. Therefore, with both EPA and Competencies, especially MK, End R1 faculty EPA/Competencies ratings carry a prospective ITE signal that is incremental to the resident’s own earlier test performance and, consistent with Q4a, independent of the most recently seen ITE result.

**Table 4.** A Early workplace-rating and baseline ITE in relation to Final ITE. Pearson correlations of End-R1 workplace ratings (six ACGME sub-competencies, the global Competency mean, and Overall EPA) with the baseline ITE (End R2) and the Final ITE. r (n).

| Outcome (ITE timepoint) | ITE End R2 | PC | MK | SBP | PBLI | PROF | ICS | All Comp | Overall EPA |
| --- | --- | --- | --- | --- | --- | --- | --- | --- | --- |
| End R2 ITE (baseline) | - | -.04 (42) | +.28 (42) | +.08 (42) | +.17 (42) | +.03 (41) | -.04 (42) | +.18 (42) | +.02 (43) |
| Final ITE | +.82*** (30) | +.15 (30) | +.44* (30) | +.25 (30) | +.45* (30) | +.37* (29) | +.16 (30) | +.31 (30) | +.49** (31) |

ITE R2 = In-Training Examination administered at the start of PGY-2. Final ITE = end-of-residency scaled score. PC = Patient Care; MK = Medical Knowledge; SBP = Systems-Based Practice; PBLI = Practice-Based Learning and Improvement; PROF = Professionalism; ICS = Interpersonal and Communication Skills; All Comp = mean of all six ACGME sub-competency ratings; Overall EPA = mean of all EPA ratings. \* p < .05; \*\* p < .01; \*\*\* p < .001.

**Table 4.** B Prospective linear regression models predicting Final ITE: End-R1 workplace ratings with ITE R2 as control.

|  | Model 5f DV: Final ITE (forced entry, n = 29) |  |  |  |  | Model 5g DV: Final ITE (forced entry, n = 30) |  |  |  |  | Model 5h (stepwise) DV: Final ITE (forward stepwise, n = 29) |  |  |  |  |
| --- | --- | --- | --- | --- | --- | --- | --- | --- | --- | --- | --- | --- | --- | --- | --- |
| Predictors | B | 95% CI | p | Tol | VIF | B | 95% CI | p | Tol | VIF | B | 95% CI | p | Tol | VIF |
| ITE R2 (control / baseline) | 0.930 | [0.65, 1.21] | <.001 | .92 | 1.09 | 0.916 | [0.67, 1.17] | <.001 | .95 | 1.05 | 0.897 | [0.64, 1.16] | <.001 | .91 | 1.10 |
| MK at End R1 | 55.01 | [3.5, 106.6] | .037 | .92 | 1.09 | - | - | - | - | - | - | - | - | - | - |
| Overall EPA at End R1 | - | - | - | - | - | 96.88 | [29.5, 164.2] | .006 | .95 | 1.05 | 98.98 | [30.33, 167.64] | .006 | .91 | 1.10 |
| Constant | B = 43.12 (SE 59.58) |  |  |  |  | B = - 44.01 (SE 70.71) |  |  |  |  | B = - 41.46 (SE 71.69) |  |  |  |  |
| Model fit | R<sup>2</sup> = .717, adj R<sup>2</sup> = .695 F(2,26) = 32.98, p < .001 N = 29 |  |  |  |  | R<sup>2</sup> = .758, adj R<sup>2</sup> = .740 F(2,27) = 42.23, p < .001 N = 30 |  |  |  |  | R<sup>2</sup> = .750, adj R<sup>2</sup> = .731 F(2,26) = 38.90, p < .001 ΔR<sup>2</sup> = .085, p = .006 N = 29 |  |  |  |  |

Dependent variable in all models: Final ITE scaled score. ITE R2 = In-Training Examination administered at start of PGY-2 (baseline knowledge control). Overall EPA = mean of all End-R1 EPA ratings; MK = Medical Knowledge mean at End R1. B = unstandardized regression coefficient; 95% CI = 95% confidence interval for B; Tol = tolerance; VIF = variance inflation factor. All VIFs ≤ 1.10, indicating negligible collinearity in all three two-predictor models.

### Q5. Blueprint fidelity

#### Sensitivity and specificity analysis

At each time point, EPAs are assessed and evaluated against the mapped competencies. The rating correlates with the Program blueprint mapping, which was studied to validate the blueprint. The observed pattern of significant EPA-by-Competency-domain correlations at End R2, the timepoint with the largest analytic sample, was compared with the official blueprint mapping. Sensitivity (the proportion of blueprint-mapped cells with a significant empirical correlation) was 18%; specificity (the proportion of unmapped cells correctly non-significant) was 71%. A 2×2 χ² test of the full 37 × 6 correlation matrix against the blueprint yielded χ²(1) = 2.5, p = 0.115; the empirical pattern did not correspond significantly to the pre-specified mapping. At End R2, only 30% of EPA × subcompetency cells reached r ≥ 0.40 (4% reached r ≥ 0.60), and content-matched pairs (e.g., EPA7 × PC2 = 0.40; EPA9 × PC1 = 0.36) were at or below threshold. By End R3, 80% of cells reached r ≥ 0.40 (37% reached r ≥ 0.60), and the same content-matched pairs strengthened to 0.60-0.81, indicating substantial blueprint structure that was masked at End R2 by insufficient between-resident variance.

Worth noting PC5 (procedural skills) showed the weakest associations across the matrix. PC5 × EPA correlations ranged from ρ = 0.12 (EPA17) to ρ = 0.64 (EPA10), strongest with EPA10 (Common procedures), EPA9 (Acute illness/injury, 0.60), EPA30 (Quality improvement and safety, 0.53), and EPA11 (Prenatal and Delivery, 0.50). With the other 18 subcompetencies, PC5 correlated poorly to average ρ = 0.35-0.66 (mean 0.50), weakest with PROF3 (0.35) and ICS2 (0.42), and, within its own PC domain, weakest with PC2 (0.43) and PC3 (0.44). Appendix 3 shows the growth trajectories of the procedural EPAs, which showed their largest gains in milestone achievement during the R4 year, with EPA10 doubling from 14% at End R3 to 42% at End R4, and EPA14 increasing from 42% to 100% over the same period. Yet by End R4, procedural EPAs remained among the lowest-attaining group: only 42% of residents reached the ≥ 3.5 threshold on EPA10 at graduation, among the lowest rates across the 37 EPAs, with only end-of-life care (EPA12, 9%), community emergency preparedness (EPA28, 29%), and prenatal/labor-and-delivery (EPA11, 36%) lower.

#### Within-domain growth-slope

This misalignment does not indicate a global impression rating process; however, the Q2 longitudinal halo analysis showed strong within-domain growth-slope correlations (r = 0.61) alongside near-zero across-domain correlations (r = 0.37), ruling out a generalized “good resident” rating effect. Taken together, the Q2 and Q5 findings are consistent with structured domain-level rater discrimination that does not map onto the specific EPA-to-subcompetency mapping prescribed by the blueprint, likely reflecting the inherently multi-domain nature of EPAs, limited statistical power with Bonferroni correction at a single time point, or imperfections in the blueprint specification itself.

#### Milestones’ competencies - EPA relation

Bringing together the findings from Q3 and Q4c characterizes how EPAs and Milestones relate. At the item level, EPA and Subcompetency correlations rose from 1 of 222 significant cells at End R1 to 18 at End R2 and 36 at End R3 (appendix 6), before softening to 19 at End R4 (most plausibly reflecting reduced power at n = 16 and range restriction as ratings compress toward Level 4). At the resident-mean level, the two instruments behaved as overlapping predictors of Final Competency: End R1 EPA was the strongest single predictor (R² = 0.420, B = 0.96, p < .001), End R1 Competency predicted more weakly (R² = 0.244, B = 0.633, p = .006), and End R2 Competency reached comparable strength to End R1 EPA six months later (R² = 0.365, B = 0.846, p < .001) tables 2, Table 4. In every model, the cross-instrument predictor at the same timepoint and baseline ITE R2 were excluded by stepwise selection (all p ≥ .32). EPAs and Milestones are therefore complementary at the level of specific content (different items measure different specific behaviors, with item-level alignment developing across training) and with the EPA mean reaching usable predictive strength earlier than the Competency mean.

## Discussion

This study provided descriptive, diagnostic, and predictive analytics of routine Clinical Competency Committee (CCC) data and explored its implications for a generalizable competency-based residency program. Our concordance and trajectory findings align with the broader EPA validation literature. Larrabee et al. first demonstrated strong concurrent EPA-Milestones correlation in pediatric residents (median R² = 0.81) [27], later replicated in pediatric fellows by Mink et al. [28], consistent with the convergence we observed by End R3. Likewise, our growth trajectories parallel the EPA developmental curves reported across 23 programs by Schumacher et al. [29] and the longitudinal milestone trajectories described in family medicine by Park et al. [21], extending both to an ACGME-I family medicine cohort.

All three instruments increased monotonically across the eight assessment time points, as expected in, and consistent with, a developmentally coherent training program. Milestones-rated competence of Al Ain four-year program graduates is comparable to that of US three-year graduates [21]. This may suggest time-based consolidation, with residents needing more time to reach US 3-year graduate levels, but another factor is the curriculum-content extension, i.e., the teaching of additional R4 content beyond the 3-year curriculum. This is supported by three findings. First, the fastest-growing EPAs between End R3 and End R4 are in areas included in the R4 curriculum: Teaching and supervising learners, Urgent/emergent care recognition, Management plans, Differential diagnosis, and Difficult conversations. Second, R4-heavy clusters showed mean changes ≥0.43 units, while the scholarship cluster, with mostly complete R3 delivery, changed by just 0.21 units. Third, three domains exceeded the US End-PGY-3 mean at End R4 (Professionalism +0.34, Practice-Based Learning +0.25, Medical Knowledge +0.14), suggesting that content extension raised ratings beyond the 3-year endpoint. Evidence from US and regional 4-year programs supports this interpretation. Carney et al. found that the 4-year advantage only manifested itself in three EPAs (comprehensive longitudinal care, prenatal care, intrapartum/postpartum care), corresponding to curricular content with additional R4 emphasis [30]. Lefevre et al. reported that a 4-year Area of Emphasis (Maternal-Child Health or Advanced Rural/Global Medical Services) provided a broader cognitive and procedural scope than 3-year or 3-year-plus-fellowships, but no improvements on the (3-year) shared curriculum [31].

This aligns with the EPAs’ entrustment framework by Ten Cate et al., entrustment ratings increase with curriculum delivery and observation, not time [32]. The other possible contributing factors are context-related or within-rater understanding. Multiple studies show rating standards are locally anchored: raters use context-specific schemas [33], programs drive most Milestone variance [34], CCCs build their own internal anchors [7], and cross-program PROF/ICS ceiling patterns reflect context-dependent rater bias rather than true ceiling [35].

The domain-specific pattern, strong gains in cognitive and professional domains, comparable performance in clinical and systems domains, is broadly consistent with US national reports [36]; [37]; [21, 38], and cross-specialty findings that clinical-procedural subcompetencies lag behind cognitive ones [39, 40].

Finally, when comparing later to earlier cohorts [41], one finds that the graduating all-competency Milestone mean rose from 3.8 to 4.00 with narrower between-resident SD, and the rating range expanded from 1.2 to an appropriate 2.4 level aligned with US entry norms [21], and the ITE benchmark improved from 343.5 (below the 380 pass standard) to 451 a 108-point gain that cannot reflect rating drift. These shifts coincide with the 2018 introduction of the 37-EPA framework, with the largest Milestone gain in MK (+0.36) paralleling the ITE improvement, consistent with the impact of CBME implementation, though causal attribution is precluded by the observational design.

### Between-cohort differences and rating-system maturation (Q1c)

The difference between cohorts on EPA ratings, but not on Competency ratings (End R3 cohort gap of only 0.04 points), is informative because it rules out genuine between-cohort differences in resident performance: the same residents in the same program show stable cross-cohort behavior on the long-established Milestones instrument and progressive improvement in cohort ratings over the years on the newly introduced EPA instrument. This pattern is consistent with rating-system maturation rather than a true cohort effect. [20] International experience supporting this interpretation includes the AAMC’s Core EPA pilot across 10 US medical schools, which took 5 years to establish reliability in entrustment, assessment, curriculum, and faculty development in assessment [42]. [33, 43].

### Rater behavior: straight-lining, discrimination, and halo (Q2)

Three findings demonstrate a disciplined assessment behavior in supervisor ratings. First, Milestones and EPA straight-lining rates were substantially lower than those reported in US adjacent specialties: pooled R1-R3 Milestones straight-lining was 2.3%, below US Emergency Medicine (∼6.0%) and Oncology (∼12.1%) [44, 45], and EPA straight-lining was 0% throughout training. The R4 rise in straight-lining (25% at Mid R4) is probably due to ceiling effects. The complete absence of EPA straight-lining likely reflects the fact that straight-lining is mathematically less likely on a 37-item instrument than on a 19-item one.

Second, between-resident SD remained narrow but stable throughout training, suggesting that supervisors continued to discriminate between residents on individual items even as the cohort matured. Third, the longitudinal halo analysis, which examines within-domain versus across-domain growth-slope correlations, produced the opposite of the halo pattern. These findings indicate that the program’s CCC engaged substantively with the rating task during the first three training years.

### Concurrent EPA-Milestones agreement (Q3)

EPA and Milestones ratings showed expected concurrent association. While a meta-analysis reported r = 0.59 between Milestones and EPA, but no link with USMLE scores or other outcomes [46], our correlations averaged r = 0.65 across eight time points, with the strongest mid-training, aligning with ranges reported in other studies [28, 47]. This suggests that EPA and Milestones provide partly overlapping, but partly complementary information. Milestones measure attainment within subcompetencies; EPAs assess integrated task performance [14, 32]

### Predictive diagnostic analytics

#### Prospective prediction of graduation outcomes (Q4c)

Predicting graduation or near-graduation (R4 or R3 if R4 not yet reached) outcomes from early competency ratings is consistent with [38], which showed that - across emergency medicine, family medicine, and internal medicine - a milestone rating of ≤ 2.5 at the end of PGY-2 predicted failure to attain the Level 4 graduation goal with probabilities of 32-67%. Our study adds an important perspective, with an earlier End R1 than the End-PGY-2 anchor used by Holmboe et al. Possible explanations are, first, the maturation of the CBME and CCC, with more than ten years of ACGME-I accreditation experience, to the point of producing discriminating ratings from the first full year of resident-rater contact. Second, EPAs and Milestones may be operating as independent, complementary instruments at the earliest training stage. Despite this early independence, End R1 overall EPA already predicted terminal Competency more strongly than any single Milestones domain at the same time point. EPAs, therefore, had real predictive ability at End R1, whereas Milestones did not at the same time point. This may not be because EPAs are a better instrument, but because the two instruments capture different aspects of resident development, [48]. The implication for programs using both instruments is that one instrument’s mean may be sufficient for resident-level decisions (selection, identification of struggling residents, summative judgment), while both instruments are needed for the formative work of feedback and targeted observation. [29]. Recently, a three-year, 48-program prospective cohort study of 4,328 pediatric residents developed and validated a multilevel structural equation model that predicts individual residents’ milestone levels from their EPA entrustment-supervision ratings, with external prediction correlations of 0.68-0.72, providing strong evidence that EPAs and Milestones carry mutually predictive, not merely concurrent, information [29]. Our finding that EPA ratings predict Competency at graduation, and predict ITE more strongly than the Competency aggregate, is consistent with this emerging literature and calls for further work to characterize the directional asymmetry across specialties and rating cultures.

#### Anchoring bias, MK validity, and the post-graduation question (Q4a, Q4b)

The finding of a lower probability of rater anchoring bias driving MK Competency as well as MK being the strongest concurrent correlate of ITE among the six Comp domains, with no other domain showing this consistency, is consistent with published Milestones validity work in family medicine [49] and other specialties [50] and reflects content alignment between the MK domain and ITE rather than a limitation of the Milestones framework. Cross-sectional correlation of medical knowledge-based assessments and milestones, and prospective prediction of ITE from early-residency milestone ratings, has been described in other specialties [51], with findings similar to ours. Medical Knowledge competency ability to predict near-graduation ITE beyond early ITE, as well as overall EPA at End R1, with EPA being a stronger predictor, highlights the value of supervisors’ early judgments. This is in line with the concept that EPA ratings capture supervisors’ impressions across multiple observations and competencies [32].

Future research is needed to test the predictive validity of Milestones/EPAs, specifically whether ratings forecast post-graduation performance. Results are mixed, with a focus on validity during training rather than on practice after graduation [52–55]. Research is needed to examine whether raters and context account for these mixed results, and whether outcomes-based benchmarks for non-knowledge skills are needed to calibrate ratings against post-graduation outcomes.

#### Blueprint fidelity and timing of measurement (Q5)

A program-level analysis of whether supervisors’ EPA ratings aligned with the program’s intended blueprint, mapping each EPA to its expected ACGME-I subcompetencies, yielded the important methodological observation that fidelity is partly dependent on measurement time, which supports the need to report detailed blueprint-fidelity statistics across multiple timepoints. Blueprint-mapped cells that remain low, for example, an EPA on chronic disease management that the blueprint maps to Medical Knowledge but that shows weak empirical correlation with MK ratings, can be interpreted as program-level signals of curriculum implementation gaps or assessment-coverage gaps rather than as evidence against the blueprint itself, since the blueprint reflects specialty-community consensus on the expected mapping. Conversely, unmapped cells with consistently high correlations may surface unintended but real competency loadings worth incorporating into local assessment design. In this framing, blueprint fidelity serves as a program-level diagnostic that translates an accreditation-approved mapping into actionable feedback on the program’s curriculum delivery and assessment culture.

Finally, the misalignment did not reflect a global-impression rating process, since the longitudinal halo analysis ruled that out. Instead, the pattern was asymmetric across the six domains: ICS and PROF subcompetencies attracted the densest EPA loadings (ICS1, ICS3, PROF3), while Patient Care and Medical Knowledge subcompetencies attracted considerably fewer EPA loadings. National-level OBGYN data show similar substantial program-level variation in milestone ratings for ICS and PROF across all PGY levels [34]. This pattern may reflect rater observation patterns rather than construct invalidity of the EPA instrument itself: communication and behaviors are directly observable in nearly every clinical encounter, whereas patient-care and medical-knowledge subcompetencies require more specific clinical content to be evident. Similar to the exact analysis, EPA and subcompetencies cross-loadings are not well described in the CBME literature and are limited only to areas such as what can affect supervisors’ entrustment in EPA, for example. Additionally, milestone ratings may reflect institutional norms rather than resident-level performance, which complicates the interpretation of rating-blueprint alignment. EPAs with the broadest multi-subcompetency loadings were EPA20 (History and physical examination, 16 subcompetencies), EPA31 (Teaching and supervising learners, 15), EPA33 (Management plans, 15), EPA35 (Differential diagnosis, 13), and EPA21 (Lab indications and interpretation, 12), all integrative clinical activities for which broad competency loading is theoretically expected. Conversely, nine EPAs (EPA5, EPA10, EPA12, EPA17, EPA23, EPA25, EPA26, EPA27, EPA30) showed no subcompetency loading above the r ≥ 0.40 threshold, likely reflecting limited R1-R2 exposure to these activities combined with small per-timepoint samples.

The Procedural skill (PC5), being distinct (not correlating well) from all other subcompetencies in the rating system, may be program-specific or specialty-specific. Program-specific issues are due to the 4-year duration of the program and procedural documentation delivered by most residents, or family medicine not being a procedurally-intensive specialty; procedures are encountered episodically. So supervisors’ longitudinal exposure to any one resident’s procedural performance is sparser than their exposure to that resident’s clinical reasoning or care coordination. Also, the scope of expected procedures in family medicine is unusually broad, ranging from joint injections to IUD insertion, suturing, and resuscitation, and entrusting this breadth may lead to a different timeline or take supervisors longer to assess confidently. This is independently supported by the pattern described of late acceleration combined with incomplete final attainment, which is consistent with procedural entrustment requiring a longer developmental window than cognitive-clinical entrustment, and possibly extending beyond the four-year training period for some residents. They are also consistent with Yamazaki et al.’s (2022) multilevel CFA finding that procedural skill is a distinct sub-factor in OBGYN PC. The practical implication is that reporting procedural skill separately gives CCCs resolution otherwise lost in a combined PC mean.

## Limitations

As a single-program study with residency programs accounting for the majority of variance in Milestone ratings nationally [11], generalisability of specific findings to other ACGME-I or ACGME-accredited programs is uncertain, though interpretation is anchored on US national norms and adjacent-specialty benchmarks. An additional limitation is that late-training Competency data rest on a single cohort, the only group with terminal data on the standardized 1-5 scale at the time of analysis, producing wide confidence intervals at R4 timepoints. Finally, non-educational factors (personality, well-being, social context) that may influence learning and assessment outcomes were not assessed.

## Conclusion

By graduation, residents demonstrated substantial and progressive competency achievement across both instruments, with the majority reaching the entrustable threshold on both EPA and Milestone ratings and ITE pass rates rising markedly across training years.

At the program level, the rating system demonstrated disciplined assessment behavior and both concurrent and prospective validity relative to the ITE. Overall EPA at End R1 emerged as the strongest prospective predictor across all three terminal outcomes, final ITE score, graduating Competency Milestones, and graduating overall EPA, outperforming both Competency Milestones and baseline knowledge as a forward-looking signal of residency achievement.

Routine CCC data support two categories of diagnostic tool. Rater-process diagnostics, covering straight-lining, within- and between-resident variation, and halo effects, provide structured feedback to raters and program leadership. Outcome-validity diagnostics, including blueprint fidelity, anchoring bias, MK-ITE correlation, and ITE prediction from workplace ratings, enable leadership to monitor criterion independence and prevent contamination. Together with the asymmetric-instrument diagnostic, these tools constitute an evidence-based quality assurance framework that requires no additional data collection beyond what programs already gather through routine CCC processes.

## Declarations

## Ethical Approval

Ethical approval was granted by the SEHA Clinics Institutional Review Board (SCIRB), ref. SCIRB/2026/16. All data were anonymized, and no consent was needed as the data were routinely gathered.

## Data Availability

The datasets generated and analyzed during the current study are not publicly available due to participant confidentiality and institutional data governance requirements.

## Authors contributions

LBK conceived the study, analyzed the data, conceptualized the quality assurance framework, and wrote the manuscript. NN reviewed the statistical analysis and the manuscript. The authors AZ and MK reviewed and approved the final manuscript.

---

**Appendix 1:**
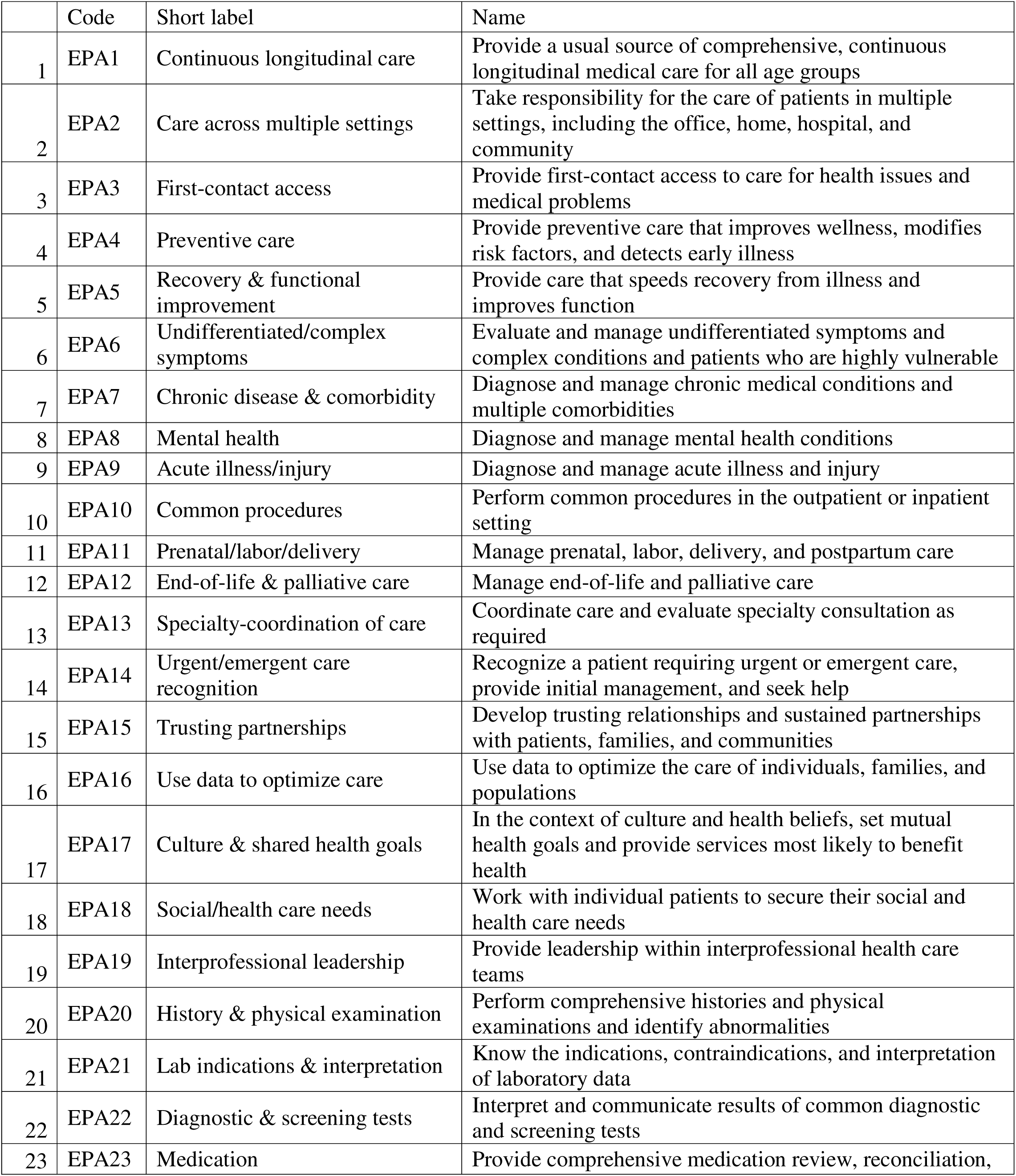
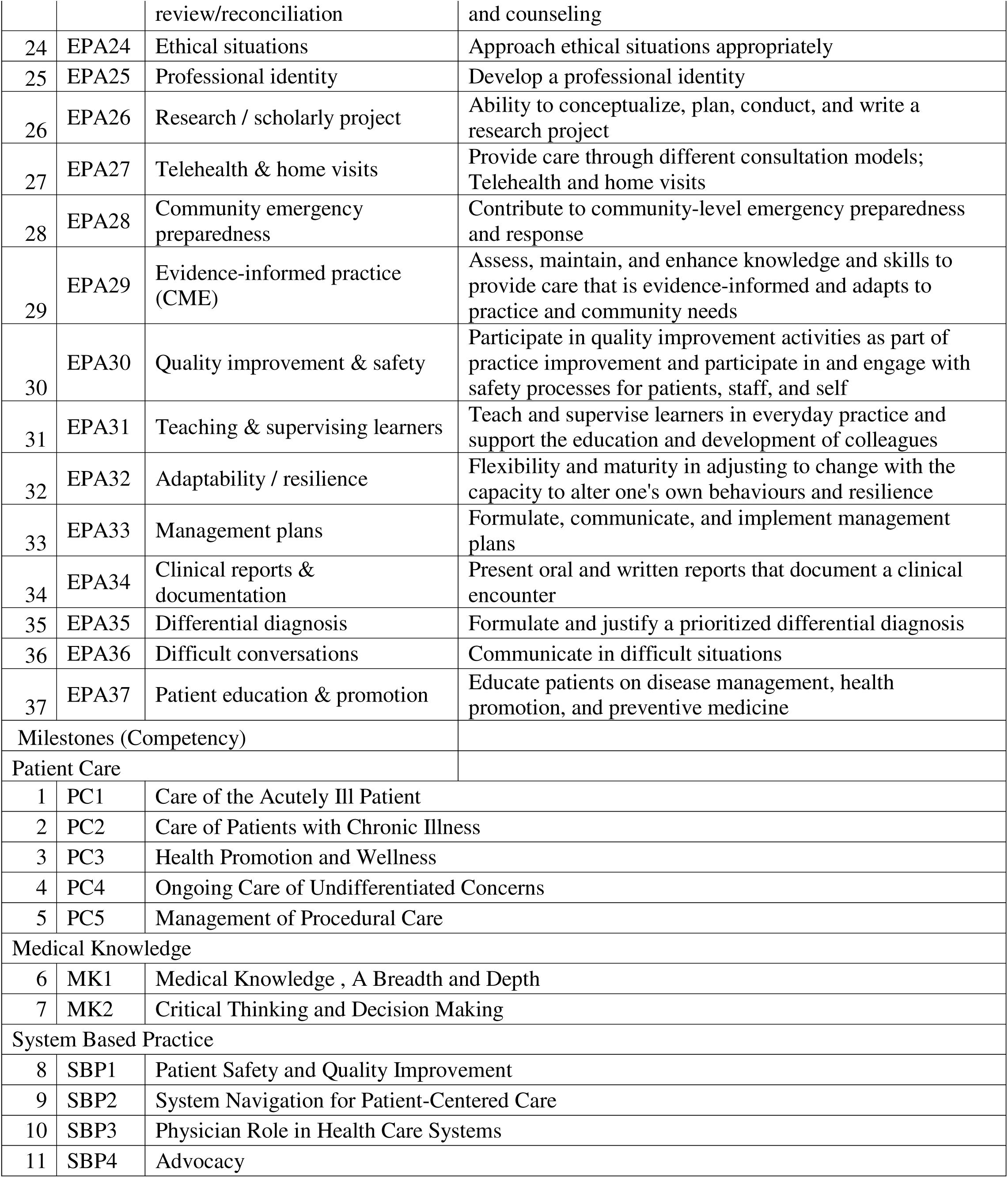
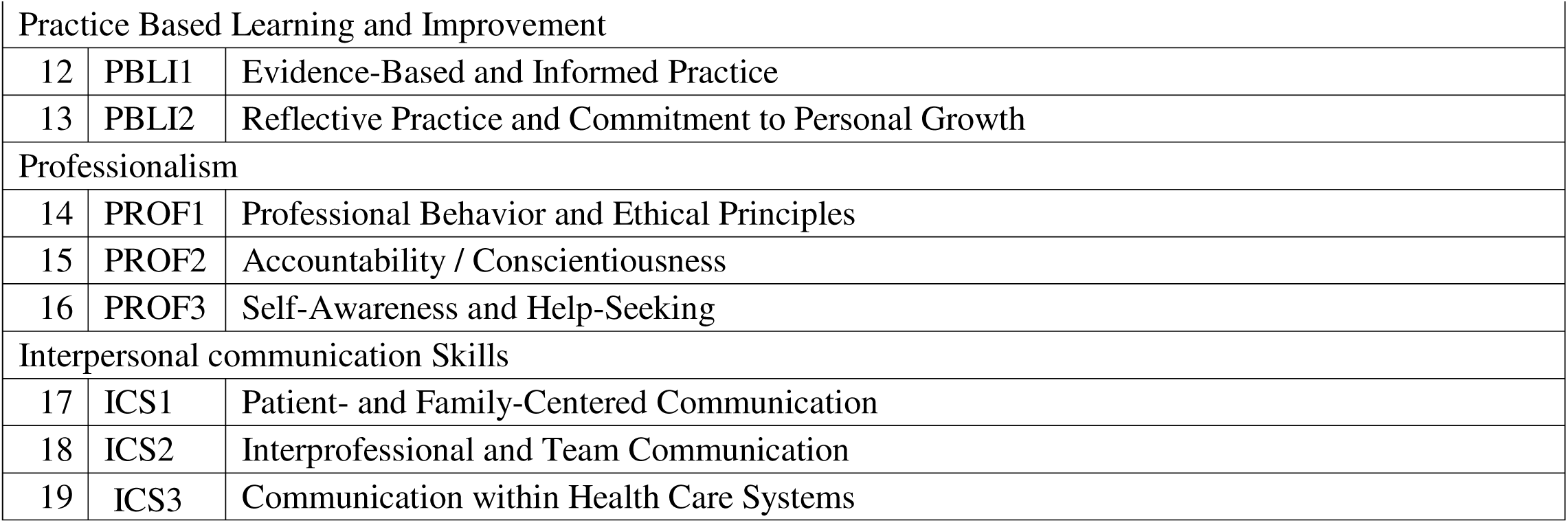

|  | Code | Short label | Name |
| --- | --- | --- | --- |
| 1 | EPA1 | Continuous longitudinal care | Provide a usual source of comprehensive, continuous longitudinal medical care for all age groups |
| 2 | EPA2 | Care across multiple settings | Take responsibility for the care of patients in multiple settings, including the office, home, hospital, and community |
| 3 | EPA3 | First-contact access | Provide first-contact access to care for health issues and medical problems |
| 4 | EPA4 | Preventive care | Provide preventive care that improves wellness, modifies risk factors, and detects early illness |
| 5 | EPA5 | Recovery & functional improvement | Provide care that speeds recovery from illness and improves function |
| 6 | EPA6 | Undifferentiated/complex symptoms | Evaluate and manage undifferentiated symptoms and complex conditions and patients who are highly vulnerable |
| 7 | EPA7 | Chronic disease & comorbidity | Diagnose and manage chronic medical conditions and multiple comorbidities |
| 8 | EPA8 | Mental health | Diagnose and manage mental health conditions |
| 9 | EPA9 | Acute illness/injury | Diagnose and manage acute illness and injury |
| 10 | EPA10 | Common procedures | Perform common procedures in the outpatient or inpatient setting |
| 11 | EPA11 | Prenatal/labor/delivery | Manage prenatal, labor, delivery, and postpartum care |
| 12 | EPA12 | End-of-life & palliative care | Manage end-of-life and palliative care |
| 13 | EPA13 | Specialty-coordination of care | Coordinate care and evaluate specialty consultation as required |
| 14 | EPA14 | Urgent/emergent care recognition | Recognize a patient requiring urgent or emergent care, provide initial management, and seek help |
| 15 | EPA15 | Trusting partnerships | Develop trusting relationships and sustained partnerships with patients, families, and communities |
| 16 | EPA16 | Use data to optimize care | Use data to optimize the care of individuals, families, and populations |
| 17 | EPA17 | Culture & shared health goals | In the context of culture and health beliefs, set mutual health goals and provide services most likely to benefit health |
| 18 | EPA18 | Social/health care needs | Work with individual patients to secure their social and health care needs |
| 19 | EPA19 | Interprofessional leadership | Provide leadership within interprofessional health care teams |
| 20 | EPA20 | History & physical examination | Perform comprehensive histories and physical examinations and identify abnormalities |
| 21 | EPA21 | Lab indications & interpretation | Know the indications, contraindications, and interpretation of laboratory data |
| 22 | EPA22 | Diagnostic & screening tests | Interpret and communicate results of common diagnostic and screening tests |
| 23 | EPA23 | Medication | Provide comprehensive medication review, reconciliation, |
|  |  | review/reconciliation | and counseling |
| 24 | EPA24 | Ethical situations | Approach ethical situations appropriately |
| 25 | EPA25 | Professional identity | Develop a professional identity |
| 26 | EPA26 | Research / scholarly project | Ability to conceptualize, plan, conduct, and write a research project |
| 27 | EPA27 | Telehealth & home visits | Provide care through different consultation models; Telehealth and home visits |
| 28 | EPA28 | Community emergency preparedness | Contribute to community-level emergency preparedness and response |
| 29 | EPA29 | Evidence-informed practice (CME) | Assess, maintain, and enhance knowledge and skills to provide care that is evidence-informed and adapts to practice and community needs |
| 30 | EPA30 | Quality improvement & safety | Participate in quality improvement activities as part of practice improvement and participate in and engage with safety processes for patients, staff, and self |
| 31 | EPA31 | Teaching & supervising learners | Teach and supervise learners in everyday practice and support the education and development of colleagues |
| 32 | EPA32 | Adaptability / resilience | Flexibility and maturity in adjusting to change with the capacity to alter one's own behaviours and resilience |
| 33 | EPA33 | Management plans | Formulate, communicate, and implement management plans |
| 34 | EPA34 | Clinical reports & documentation | Present oral and written reports that document a clinical encounter |
| 35 | EPA35 | Differential diagnosis | Formulate and justify a prioritized differential diagnosis |
| 36 | EPA36 | Difficult conversations | Communicate in difficult situations |
| 37 | EPA37 | Patient education & promotion | Educate patients on disease management, health promotion, and preventive medicine |
| Milestones (Competency) |  |  |  |
| Patient Care |  |  |  |
| 1 | PC1 | Care of the Acutely Ill Patient |  |
| 2 | PC2 | Care of Patients with Chronic Illness |  |
| 3 | PC3 | Health Promotion and Wellness |  |
| 4 | PC4 | Ongoing Care of Undifferentiated Concerns |  |
| 5 | PC5 | Management of Procedural Care |  |
| Medical Knowledge |  |  |  |
| 6 | MK1 | Medical Knowledge , A Breadth and Depth |  |
| 7 | MK2 | Critical Thinking and Decision Making |  |
| System Based Practice |  |  |  |
| 8 | SBP1 | Patient Safety and Quality Improvement |  |
| 9 | SBP2 | System Navigation for Patient-Centered Care |  |
| 10 | SBP3 | Physician Role in Health Care Systems |  |
| 11 | SBP4 | Advocacy |  |

|  |  |  |
| --- | --- | --- |
| Practice Based Learning and Improvement |  |  |
| 12 | PBLI1 | Evidence-Based and Informed Practice |
| 13 | PBLI2 | Reflective Practice and Commitment to Personal Growth |
| Professionalism |  |  |
| 14 | PROF1 | Professional Behavior and Ethical Principles |
| 15 | PROF2 | Accountability / Conscientiousness |
| 16 | PROF3 | Self-Awareness and Help-Seeking |
| Interpersonal communication Skills |  |  |
| 17 | ICS1 | Patient- and Family-Centered Communication |
| 18 | ICS2 | Interprofessional and Team Communication |
| 19 | ICS3 | Communication within Health Care Systems |

**Appendix 2.** A two-tier quality-assurance toolkit for competency-based assessment programs.

| TIER 1 - Individual-rater diagnostics (for individual rater feedback and action) |  |  |  |  |
| --- | --- | --- | --- | --- |
| Diagnostic | What it detects | Data required | Method | Threshold / flag |
| **Straight-lining rate** | Rater fatigue or disengagement, a resident receives the same rating on every item at a single timepoint | Per-resident per-timepoint item-level ratings; one row per (resident, TP) | % of residents with item-level SD = 0 at each TP | Rate > 10% at any timepoint; or any monotonic upward trend across training years (suggests worsening rater fatigue) |
| **Within-resident variation (item-level)** | Whether raters discriminate between items for a given resident, or apply a flat rating | Per-resident per-timepoint item-level ratings on one or both instruments | Compute SD across items within each resident at each TP; track the median across timepoints | Monotonic decline in within-resident SD across training (suggests increasing halo); minor decline expected as residents progress |
| **Between-resident variation (item-level)** | Whether raters discriminate between residents on items, or rate cohorts uniformly | Per-resident per-timepoint item-level ratings; pooled across residents | Compute SD across residents for each item at each TP; track the median across items | Between-resident SD < 0.30 or progressive decline across training is concerning |
| **Longitudinal halo detection** | A global 'good resident' impression that yokes scores across unrelated competency domains | Per-resident growth slopes (from linear or polynomial fits across the 8 TPs) | Compute mean Pearson r of growth slopes within ACGME core competency domains vs across-domain r | Across-domain r > 0.40 with within-domain r similar (gap < 0.20) suggests halo. Within-domain r > across-domain r is healthy. |
| TIER 2 - Program-level diagnostics (for aggregate feedback and program-level action - not for individual rater review) |  |  |  |  |
| Diagnostic | What it detects | Data required | Method | Threshold / flag |
| **Blueprint fidelity (EPA × subcompetency)** | Whether supervisors' EPA ratings empirically load onto the subcompetencies the program blueprint expects them to reflect | Same-timepoint EPA and subcompetency-level ratings per resident at the recommended timepoint (see method); plus the program's intended blueprint mapping | Compute the EPA × subcompetency Pearson correlation matrix at the FIRST timepoint where (a) residents have meaningfully differentiated AND (b) median per-cell N is adequate (typically End R3 in 4-year programs). The strict tier scheme: $r \geq 0.40$ = acceptable ($R^2 \geq 16\%$); $r \geq 0.60$ = Strong ($R^2 \geq 36\%$); $r \geq 0.80$ = Excellent ($R^2 \geq 64\%$) (Stevens 1992; Hair 2017). Report % of cells reaching each threshold and content-matched pair correlations. | < 50% of cells reaching $r \geq 0.40$ at the recommended timepoint suggests poor blueprint fidelity. Content-matched pairs (e.g., chronic-disease EPA × chronic-illness subcompetency) failing to reach $r \geq 0.40$ is a more specific indicator of fidelity failure than a low global statistic. |
| **Anchoring-bias diagnostic (MK vs prior/future ITE)** | Whether supervisors' MK ratings are anchored on recently viewed ITE results, biasing concurrent correlations upward | MK ratings at each TP + prior-year ITE + future-year ITE | Spearman $\rho$ at each TP for (MK × prior ITE) vs (MK × future ITE); compare which is stronger | $\rho(\text{prior})$ consistently > $\rho(\text{future})$ at TPs where supervisors have seen the prior ITE suggests anchoring |
| **Same-year MK ↔ ITE concurrent correlation** | Criterion validity of the MK domain against an objective knowledge measure | MK rating at each TP + same-year ITE scaled score | Pearson r of MK × same-year ITE at each training year (R1–R4) | $r < 0.30$ at any training year warrants review of MK rating practices |
| **Prospective workplace → ITE prediction** | Whether early workplace ratings carry forward-looking predictive information beyond what is in earlier ITE | End-R1 workplace ratings + ITE R2 (control) + final ITE (near-graduation, max of R3 and R4) | Multiple linear regression: final ITE ~ ITE R2 + early-training rating | Workplace predictor $\beta \approx 0$ or non-significant suggests ratings carry no incremental predictive information |

The toolkit is intended for periodic program-level review, not for evaluation of individual raters at single time points. Thresholds are suggested starting points; programs should refine thresholds based on their own rating cultures and historical data. The two-tier structure preserves outcome-validity diagnostics from rater feedback while still providing programs with actionable process-level information about rater behavior. The blueprint-fidelity diagnostic requires core programs to compute it only at developmentally meaningful timepoints (when residents have differentiated and an adequate sample size is available), as earlier timepoints will systematically underestimate true blueprint structure.

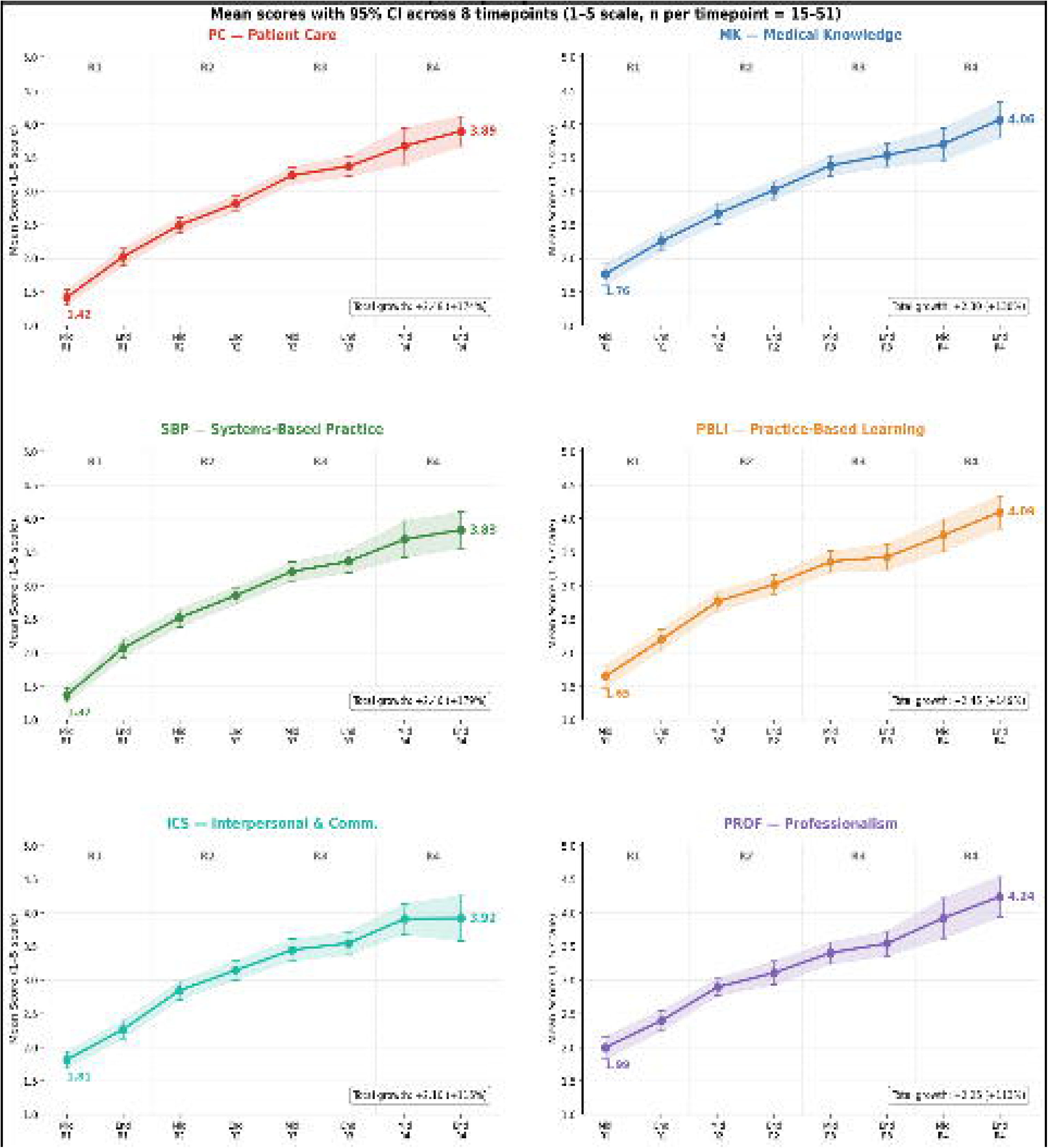

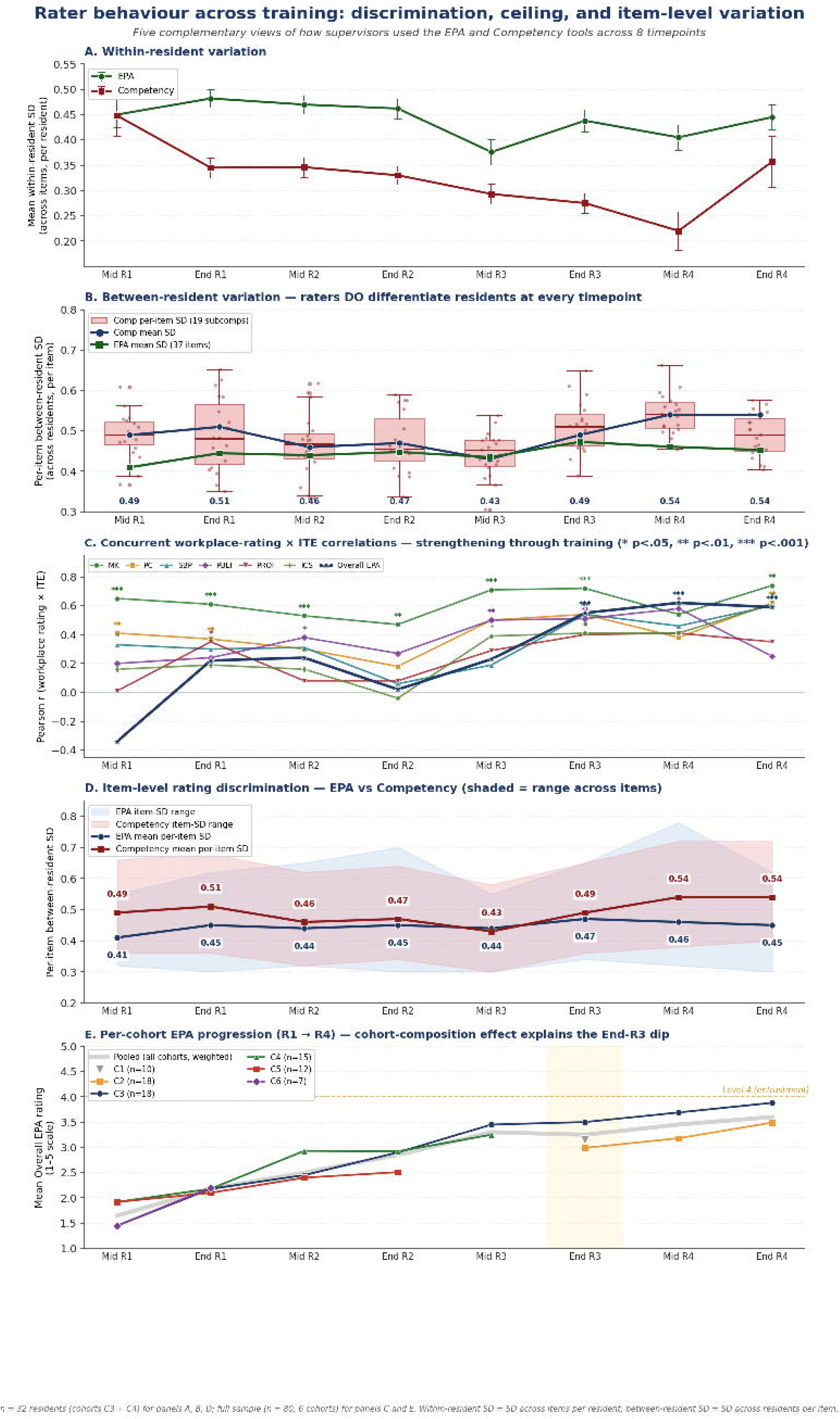

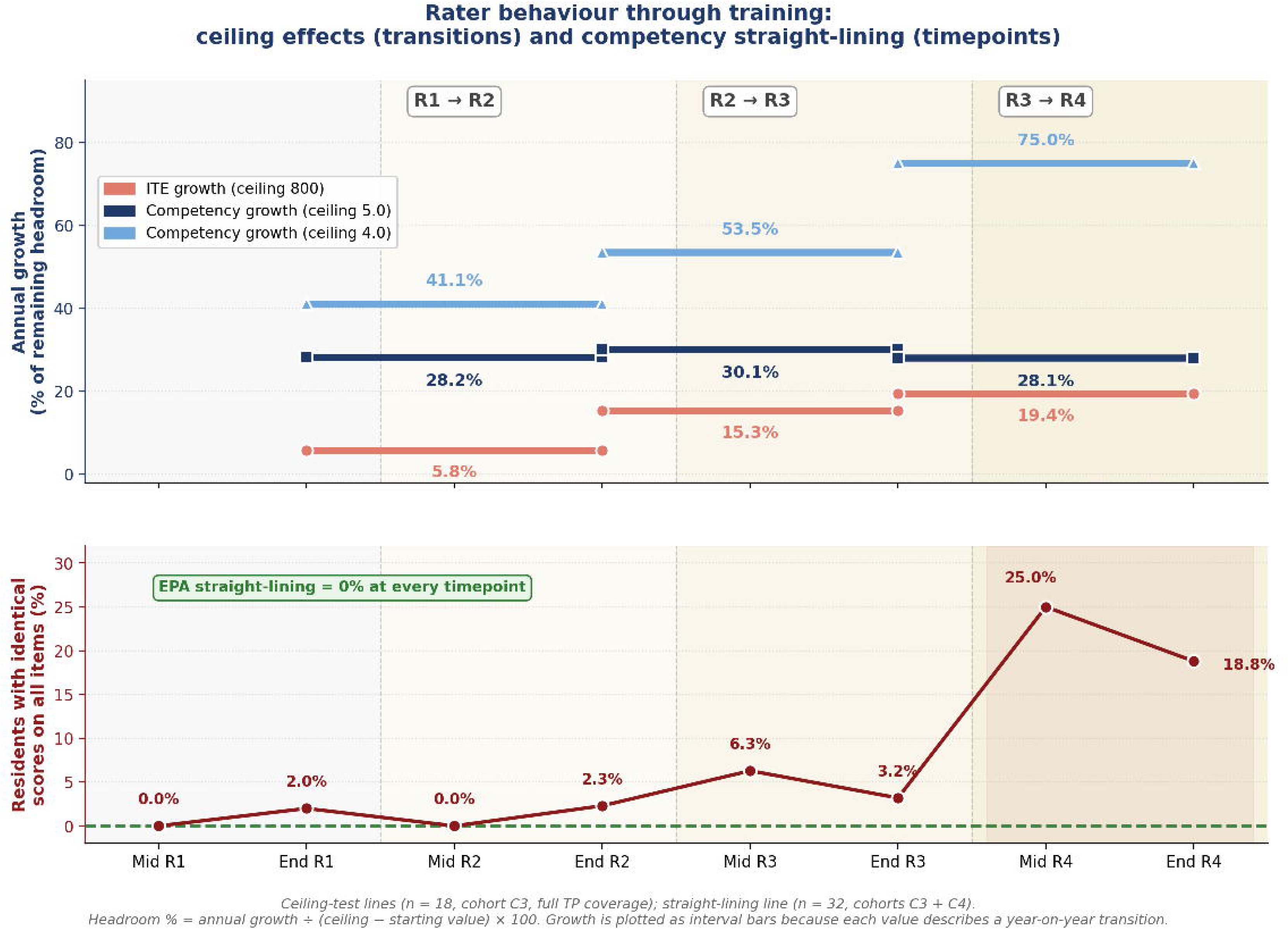

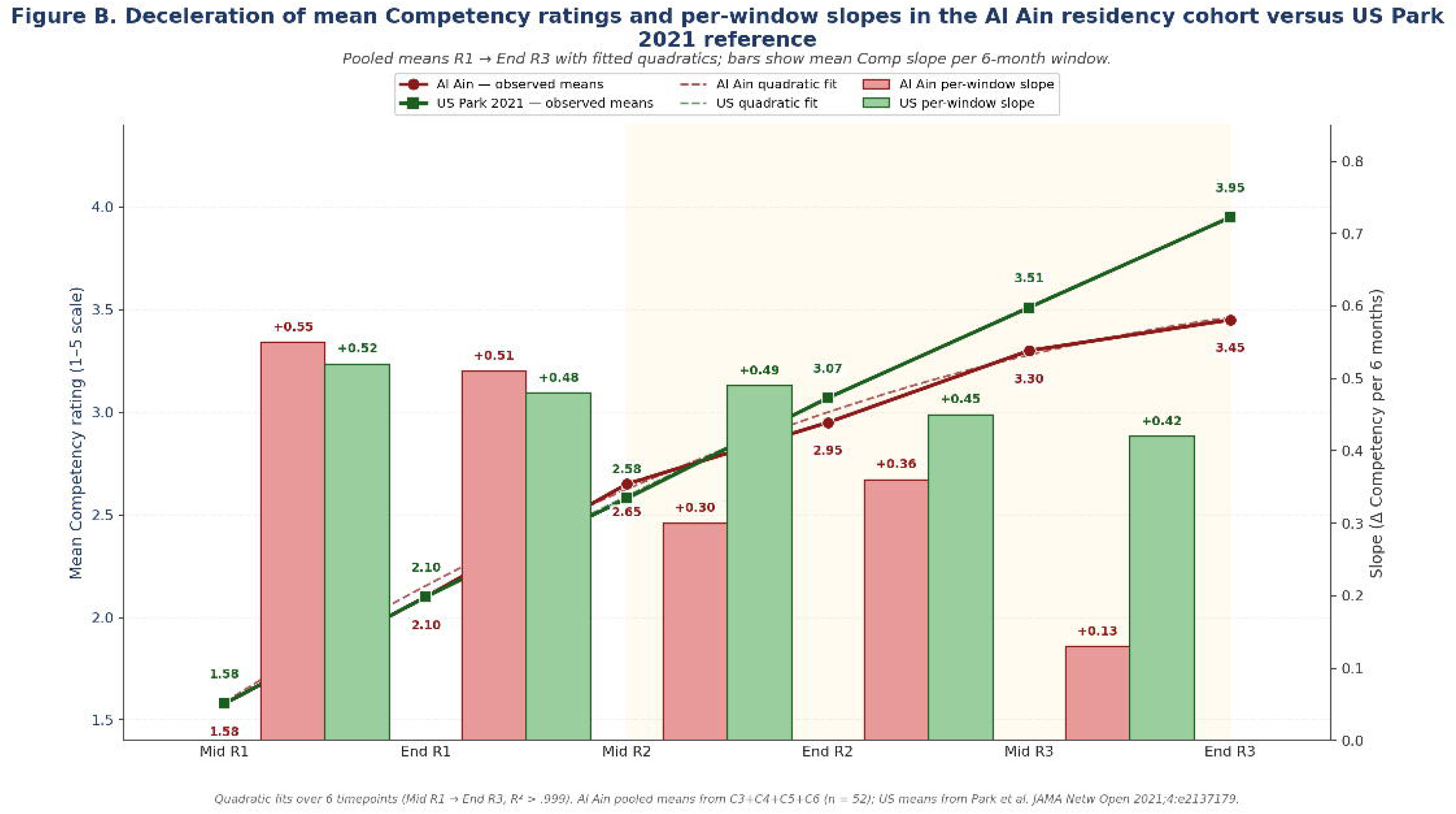

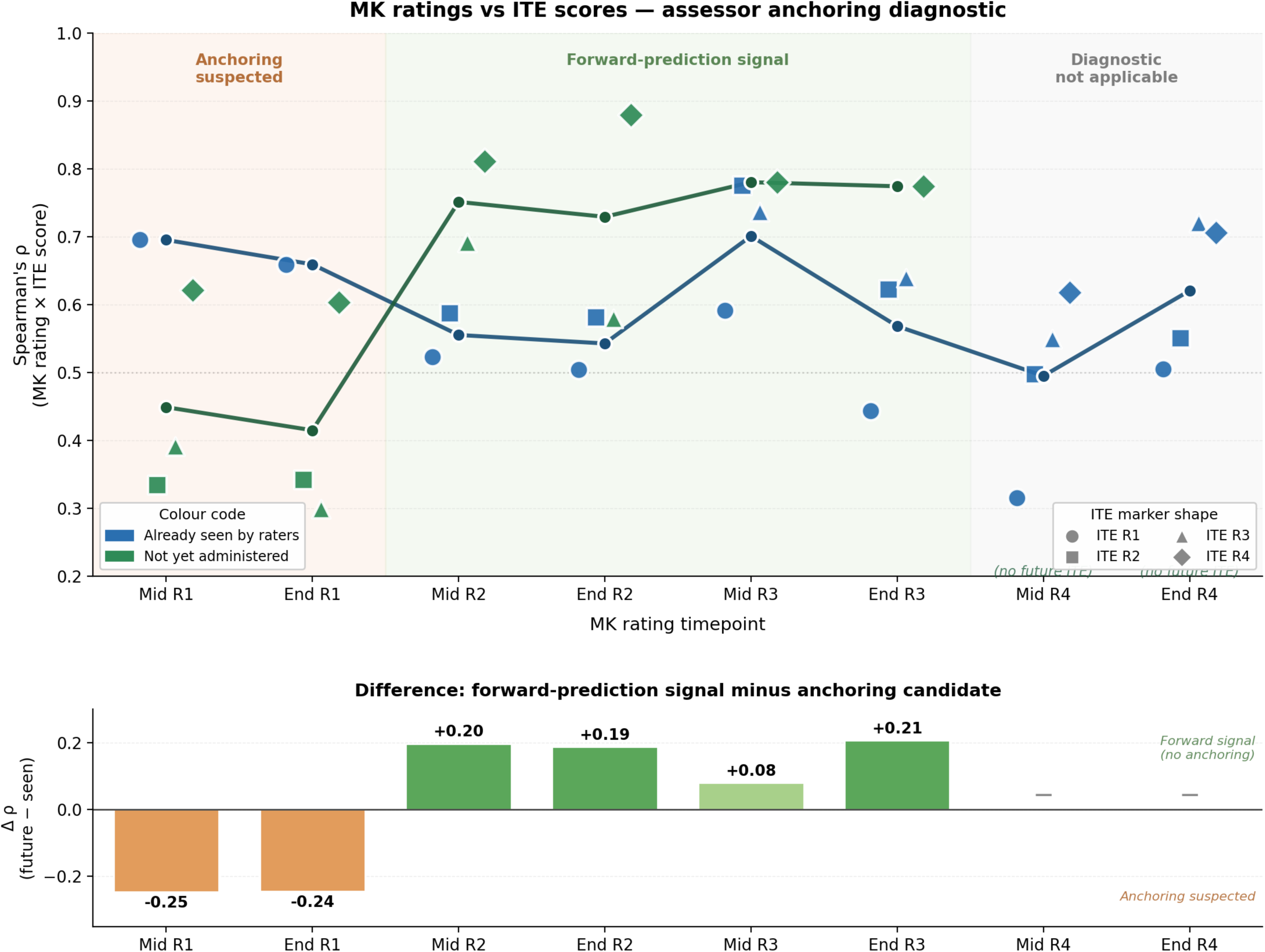

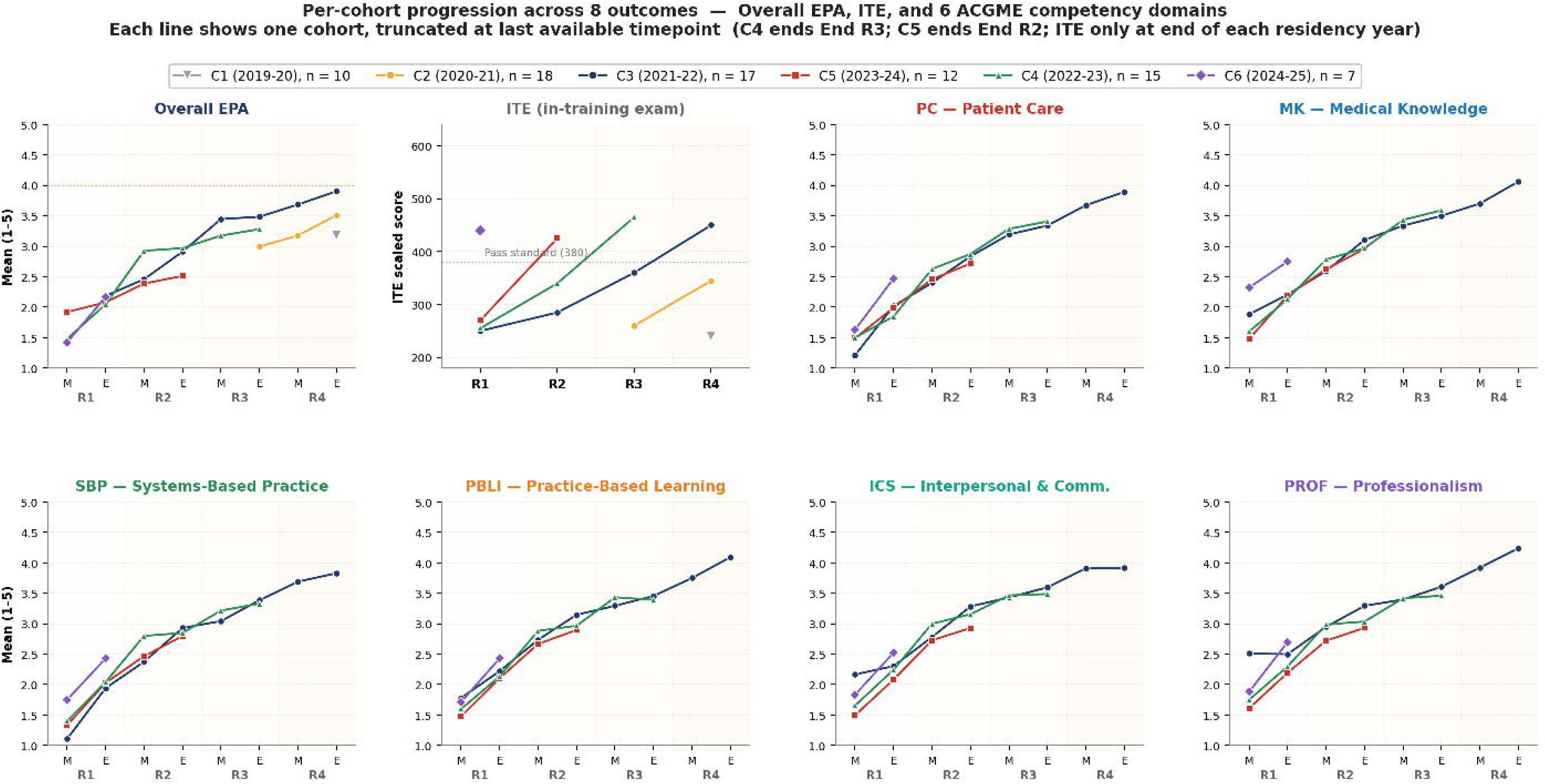

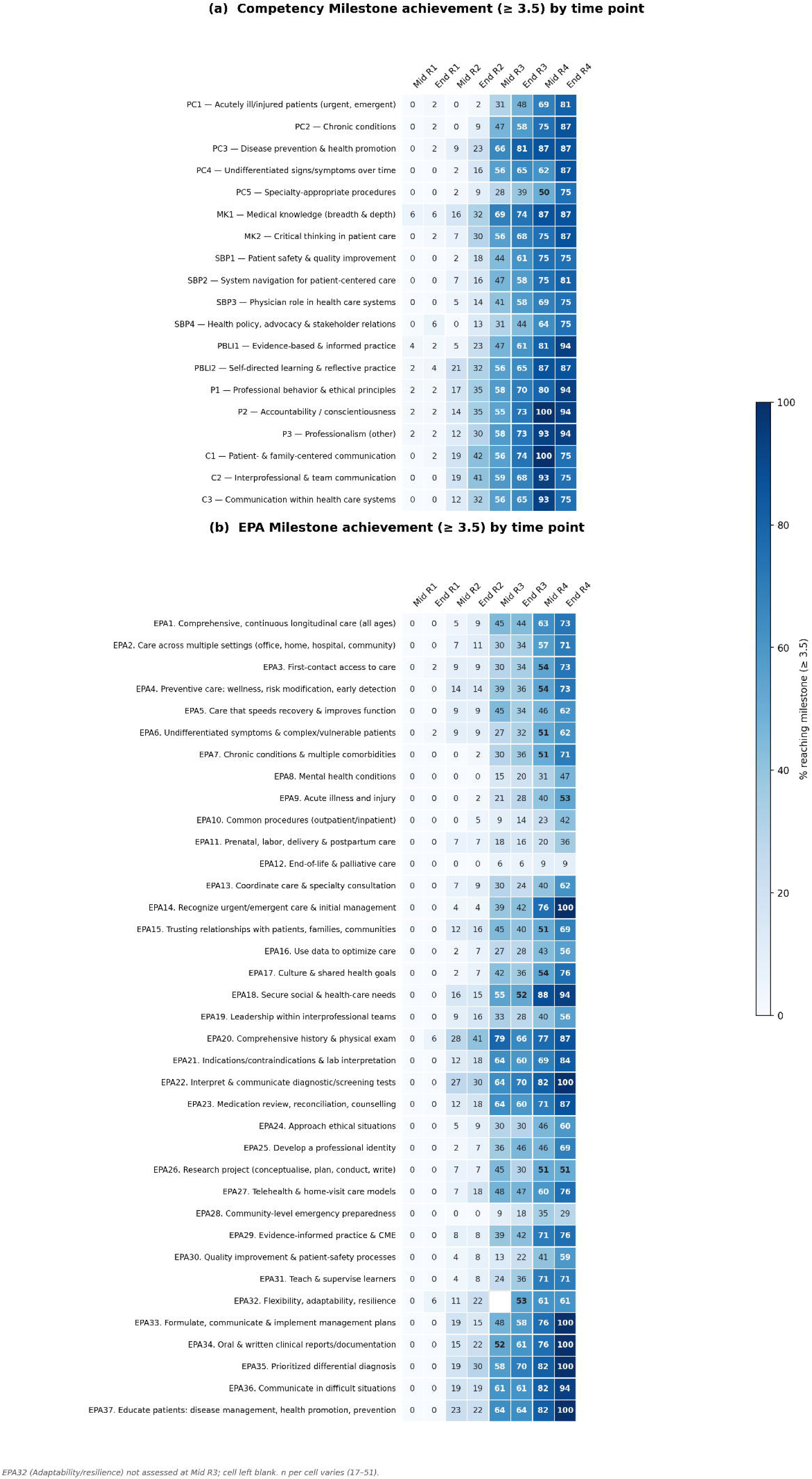

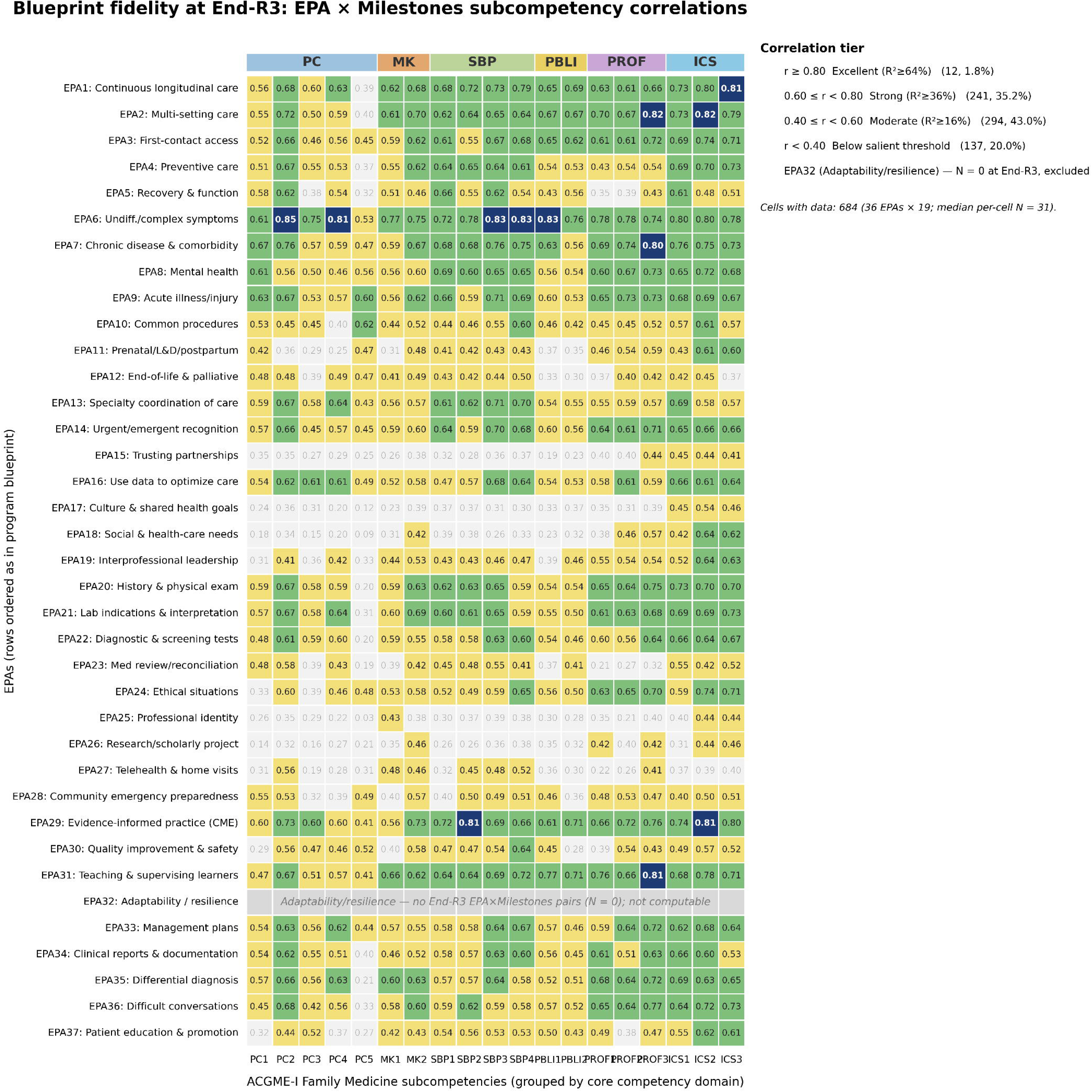

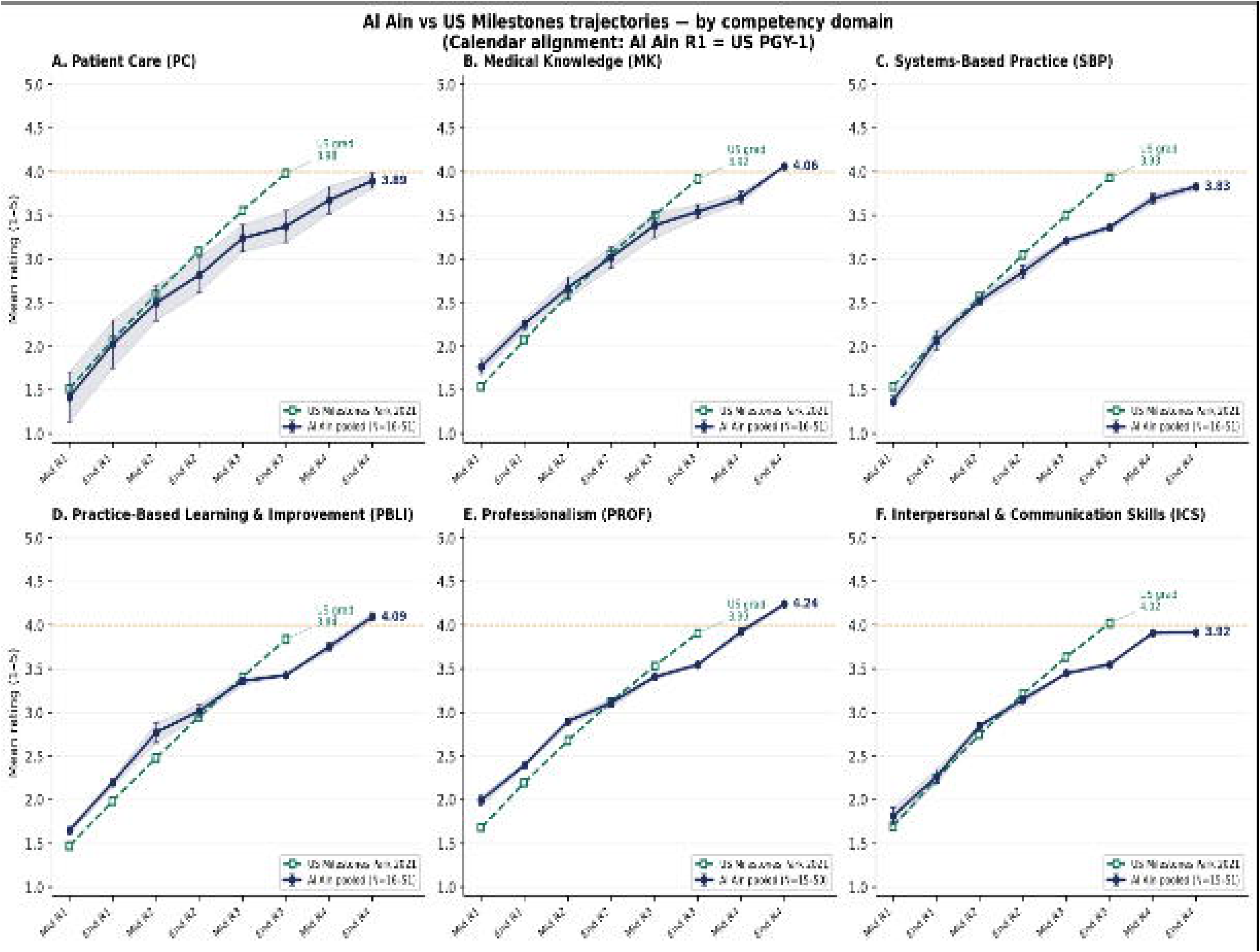

